# Human genetics implicate thromboembolism in the pathogenesis of long COVID in individuals of European ancestry

**DOI:** 10.1101/2024.05.17.24307553

**Authors:** Art Schuermans, Andreas Verstraete, Vilma Lammi, Tomoko Nakanishi, Maddalena Ardissino, Jef Van den Eynde, Benjamin B. Sun, Marios K. Georgakis, Beatriz Guillen-Guio, Louise V. Wain, Christopher E. Brightling, PHOSP-COVID Collaborative Group, Johan Van Weyenbergh, Adam J. Lewandowski, Betty Raman, Hugo Zeberg, Hanna M. Ollila, Stephen Burgess, Pradeep Natarajan, Michael C. Honigberg, Kathleen Freson, Thomas Vanassche, Peter Verhamme

## Abstract

SARS-CoV-2 infection can result in long COVID, characterized by post-acute symptoms from multiple organs. Current hypotheses on mechanisms underlying long COVID include persistent inflammation and thromboembolism; however, compelling evidence from humans is limited and causal associations remain unclear. Here, we tested the association of thromboembolism-related genetic variants with long COVID in the Long COVID Host Genetics Initiative (*n*_cases_=3,018; *n*_controls_=994,582). Primary analyses revealed that each unit increase in the log-odds of genetically predicted venous thromboembolism risk was associated with 1.21-fold odds of long COVID (95%CI: 1.08-1.35; *P*=1.2×10^-3^). This association was independent of acute COVID-19 severity, robust across genetic instruments and methods, and replicated in external datasets for both venous thromboembolism and long COVID. Downstream analyses using gene-specific instruments, along with protein and gene expression data, suggested the protease-activated receptor 1 (PAR-1) as a potential molecular contributor to long COVID. These findings provide human genetic evidence implicating thromboembolism in long COVID pathogenesis.

## Main

Infection with SARS-CoV-2 can lead to different acute clinical manifestations, ranging from mild respiratory disease to multi-organ dysfunction. It is becoming increasingly clear that infection with SARS-CoV-2 also has chronic effects, with epidemiological data suggesting that 5-10% of acutely infected individuals experience persistent complaints three months after infection^1^. These post-acute symptoms—referred to as “long COVID”—are not limited to the respiratory tract but involve various organ systems^2^. Common manifestations include physical (e.g., fatigue, post-exertional malaise, and shortness of breath), cognitive (e.g., memory loss and cognitive impairment), and psychological (e.g., anxiety and depression) symptoms^3^. Up to 85% of long COVID patients report ongoing complaints even after one year^4,5^. Despite its high prevalence and impact on quality of life, clinical strategies for the prevention and treatment of long COVID remain limited.

The lack of disease-specific therapies for long COVID can be partially ascribed to an incomplete understanding of its mechanistic drivers. Currently hypothesized mechanisms underlying long COVID include immune dysregulation, virus-induced inflammation, and endothelial dysfunction^3^. For instance, in a prospective cohort study of 1,837 adults hospitalized with COVID-19, selected blood markers of thromboinflammation (i.e., fibrinogen and D-dimer relative to C-reactive protein) predicted cognitive defects at 6-12 months after infection^6^. Another recent study of 113 individuals infected with SARS-CoV-2 revealed that the development of long COVID was associated with changes to blood proteins indicative of persistent complement activation and altered coagulation^7^. These primarily observational findings have prompted some physicians to offer treatments like apheresis and anticoagulation therapy to long COVID patients^8^. However, the evidence supporting these treatments is limited, and it remains unclear whether thromboinflammation plays a causal role in the development of long COVID.

Large-scale genome-wide association studies (GWAS) have identified numerous genetic variants associated with human diseases^9^, facilitating the identification of causal mechanisms using Mendelian randomization (MR)^10,11^. MR is an epidemiological method that uses the random assortment of alleles at conception, which leads to an effective randomization of individuals to different levels of genetic predisposition for specific traits or conditions^12,13^. This randomization limits the risk of bias due to confounding and reverse causation and can therefore provide support for a causal relationship between an exposure and outcome of interest^10,11^. Previous MR analyses have identified causal risk factors (e.g., body mass index) and molecular players (e.g., the interleukin-6 receptor) for acute COVID-19^14–16^, with clinical trial data confirming some of these as effective therapeutic targets in human patients^17,18^. Given the limitations and high costs associated with traditional methods for identifying causal mediators of diseases, genetic approaches such as MR may also help prioritize new therapeutic targets for long COVID^19^.

Here, we used MR to test the hypothesis that thromboembolism contributes causally to long COVID. In primary analyses, we constructed a genetic instrument for venous thromboembolism and estimated its causal effect on the development of long COVID. We assessed the specificity of this association and tested its robustness using different methodological approaches and external datasets. Finally, we performed gene-focused analyses to identify potential molecular mediators of long COVID and gain further insights into the possible biological mechanisms driving this condition.

## Results

### Overall analysis approach

The study design is shown in **Fig. 1**. In brief, we used a two-sample MR approach to test the association of thromboembolism-related genetic variants with long COVID. Two-sample MR leverages genetic data from two separate GWAS: the first one is used to construct a genetic instrument for the exposure of interest (e.g., venous thromboembolism), while the second is used to test the association of this instrument with the outcome of interest (e.g., long COVID)^10,20^. When an MR analysis adheres to specific assumptions (**Fig. 1**; see **Methods**), it provides a robust framework for testing the causal association of a given exposure with the corresponding outcome^10,20^.

**Figure 1.**
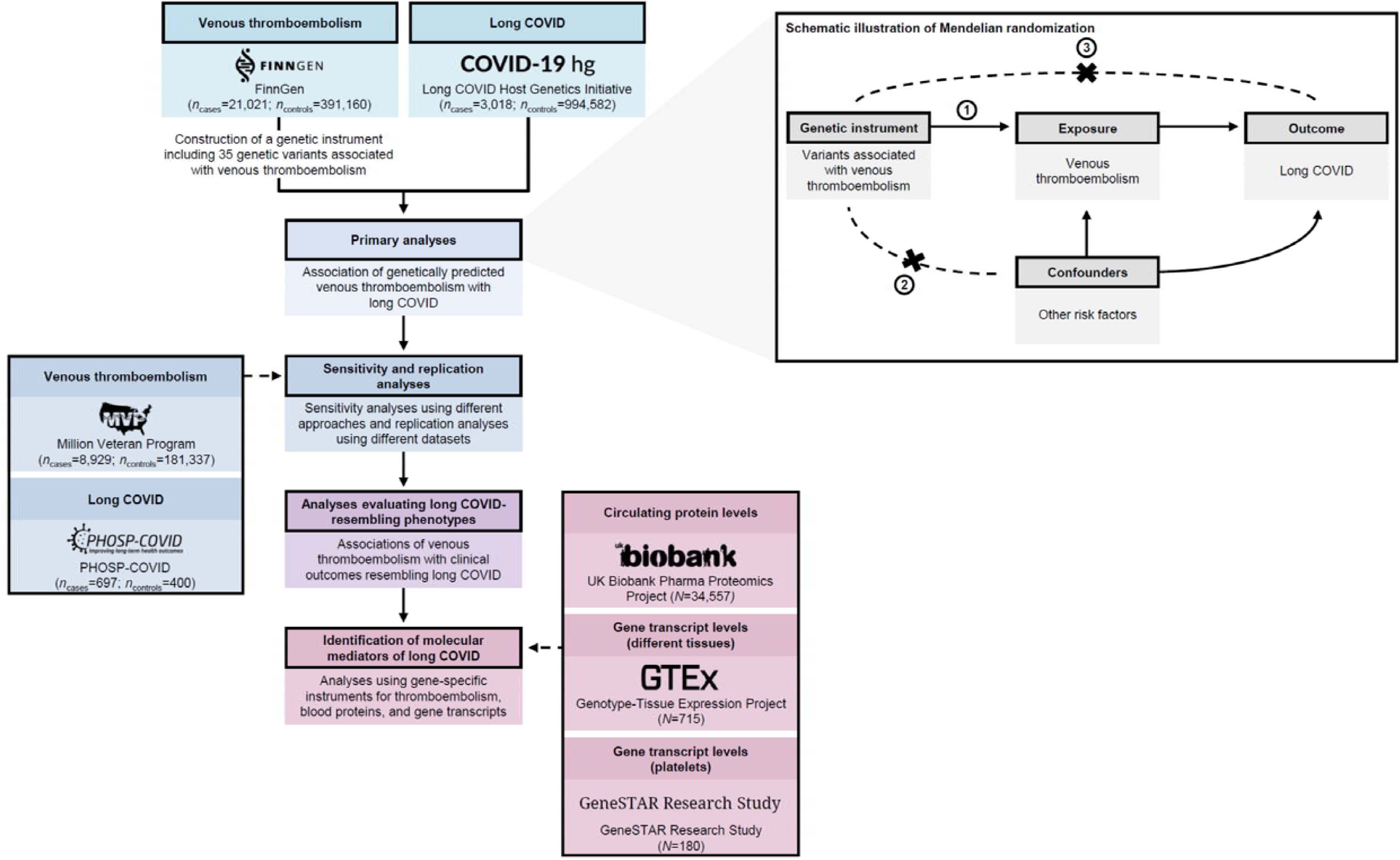
Study design. The flow chart shows the study design and the data sources. The diagram illustrates the design and three core assumptions of Mendelian randomization analyses: (1) the tested genetic instrument must be associated with the exposure of interest; (2) there must be no confounders affecting the association between the tested genetic instrument and outcome of interest; (3) the genetic instrument can only affect the outcome through its effect on the exposure of interest (i.e., no horizontal pleiotropy).

In line with this framework, we first identified 35 genetic variants associated with venous thromboembolism in FinnGen. These variants were then combined into a single statistical entity, i.e., a genetic instrument. Primary analyses tested the association of this genetic instrument (referred to as “genetically predicted venous thromboembolism risk”) with long COVID in the Long COVID Host Genetics Initiative. Downstream analyses (1) assessed the robustness of this association using different methods and external datasets; (2) tested the effects of genetically predicted venous thromboembolism risk on specific conditions with symptom profiles shared with long COVID (referred to as “long COVID-resembling phenotypes”); and (3) explored molecular mediators of long COVID using gene-focused MR approaches (**Fig. 1**).

### Cohort characteristics

The main genetic instrument for venous thromboembolism was constructed in FinnGen^21^, using data from 412,181 participants (venous thromboembolism: *n*_cases_=21,021 and *n*_controls_=391,160) from ten contributing Finnish biobanks. Venous thromboembolism cases were identified using diagnostic codes from hospital discharge and death cause registers (see **Methods**). Of the included participants, 44.1% were male, and the median age was approximately 62.9 years^22,23^.

The association of genetically predicted venous thromboembolism risk with long COVID was tested in the Long COVID Host Genetics Initiative^24^, characteristics of which are provided in **Supplementary Table 1**. While the Long COVID Host Genetics Initiative included 25 cohorts (long COVID: *n*_cases_=6,450 and *n*_controls_=1,093,995), primary analyses excluded participants who met the criteria for long COVID but did not have a prior test-verified SARS-CoV-2 infection and therefore only used data from 12 cohorts (*n*_cases_=3,018; *n*_controls_=994,582). Of these 12 cohorts, the majority (83.3%) used questionnaires rather than electronic health data to ascertain long COVID (**Supplementary Table 1**). Almost all participants (99.1%) included in these cohorts had European ancestry, 48.0% were male, and the weighted mean age across cohorts was 62.3 years^24^. Around 30.5% of long COVID cases had a history of hospitalization due to COVID-19.

### Thromboembolism-promoting genetic variants increase long COVID risk

Our main genetic instrument included 35 independent genetic variants (linkage disequilibrium *R*^2^<0.001) strongly associated with venous thromboembolism (*P*<5×10^-8^) in FinnGen (**Supplementary Table 2**). The mean *F*-statistic was 114.5, suggesting adequate strength^10^. In primary analyses, we tested the effect of this genetic instrument on long COVID risk in the Long COVID Host Genetics Initiative using the inverse-variance weighted MR method. Each unit increase in the log-odds of genetically predicted venous thromboembolism risk was associated with 1.21-fold odds of developing long COVID (95% confidence interval [CI]: 1.08-1.35; *P*=1.2×10^-3^; **Fig. 2**), indicating a significant positive association of thromboembolism-promoting genetic variants with long COVID.

**Figure 2.**
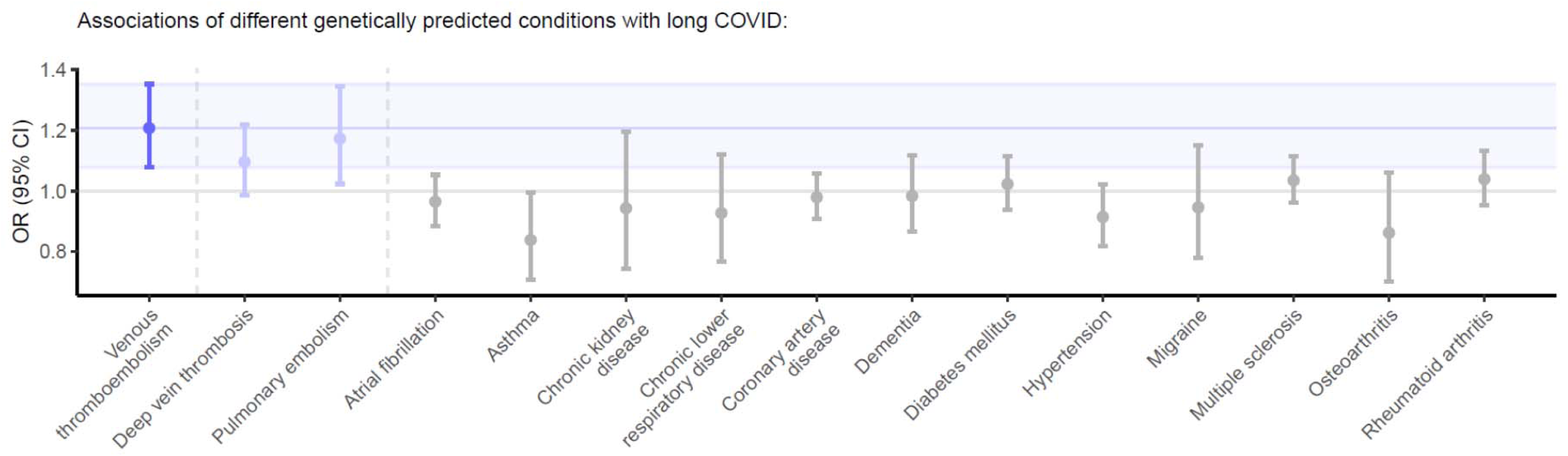
Analyses testing the associations of genetically predicted venous thromboembolism and other conditions with long COVID. Analyses testing the associations of genetically predicted venous thromboembolism and related (i.e., pulmonary embolism or deep vein thrombosis) or unrelated conditions (i.e., “negative controls”) with long COVID. The *x*-axis highlights the different conditions for which genetic instruments were constructed and associations with long COVID were tested (i.e., different exposures). All genetic instruments for the indicated conditions were constructed using genetic variants that were strongly associated with the corresponding condition (*P*<5×10^-8^) and not in linkage disequilibrium with each other (linkage disequilibrium *R*^2^<0.001) in up to 412,181 participants from FinnGen. Data for long COVID were obtained from 997,700 participants from the Long COVID Host Genetics Initiative^24^. All associations were calculated using the inverse-variance weighted method, visualized using odds ratios (ORs) and 95% confidence intervals (CIs). Genetically predicted venous thromboembolism was significantly associated with a higher risk of long COVID (depicted in dark blue; the blue band corresponds to the 95% confidence interval of this association).

To contextualize these findings, we assessed whether genetic instruments for other conditions were associated with long COVID using a similar approach. First, to corroborate the observed association of venous thromboembolism with long COVID, we tested the effects of genetic instruments for the most common manifestations of venous thromboembolism—deep vein thrombosis and pulmonary embolism—on long COVID risk. Next, to evaluate the specificity of these associations, we extended our analysis to a range of conditions representing different organ systems and pathophysiological pathways (e.g., type 2 diabetes, dementia, rheumatoid arthritis). These analyses showed that only genetically predicted risk for deep vein thrombosis (odds ratio [OR], 1.10 [95%CI, 0.98-1.22] per log-odds increase in genetically predicted deep vein thrombosis risk; *P*=9.3×10^-2^) and pulmonary embolism (OR, 1.17 [95%CI, 1.02-1.34] per log-odds increase in genetically predicted pulmonary embolism risk; *P*=2.2×10^-2^) had effect estimates within the 95%CI of the venous thromboembolism-long COVID association. None of the conditions less closely related to venous thromboembolism had effect estimates within this 95%CI or a positive association reaching nominal significance (**Fig. 2** and **Supplementary Table 3**). These findings support a specific association of venous thromboembolism with long COVID rather than a generalized association across pathophysiologically distinct conditions.

### Sensitivity and replication analyses

To evaluate the potential role of horizontal pleiotropy (i.e., effects of a genetic instrument on the outcome outside of its effects on the exposure), we tested the association of genetically predicted venous thromboembolism risk with long COVID using MR-Egger^25^. MR-Egger showed no evidence of horizontal pleiotropy (*P*_intercept_>0.99), with an effect estimate closely aligned with our primary analysis (OR, 1.21 [95%CI, 1.00-1.46] per log-odds increase in genetically predicted venous thromboembolism risk; *P*=5.1×10^-2^). This suggests that genetic variants associated with venous thromboembolism affect long COVID risk via thromboembolic mechanisms rather than alternative pathways. Consistent findings were observed with the weighted median (OR, 1.26 [95%CI, 1.06-1.49] per log-odds; *P*=7.4×10^-3^) and mode-based (OR, 1.33 [95%CI, 1.12-1.56] per log-odds; *P*=8.5×10^-4^) estimators, providing additional support for a pleiotropy-robust association.

We also performed sensitivity analyses using different linkage disequilibrium *R*^2^ (*R*^2^<0.001/*R*^2^<0.001/*R*^2^<0.01/*R*^2^<0.1) and *P*-value thresholds (*P*<5×10^-4^/*P*<5×10^-6^/*P*<5×10^-8^/*P*<5×10^-10^) to construct our genetic instrument for venous thromboembolism. These analyses showed that the association of venous thromboembolism with long COVID was directionally consistent across all tested parameters (**Fig. 3**; **Supplementary Table 4**).

**Figure 3.**
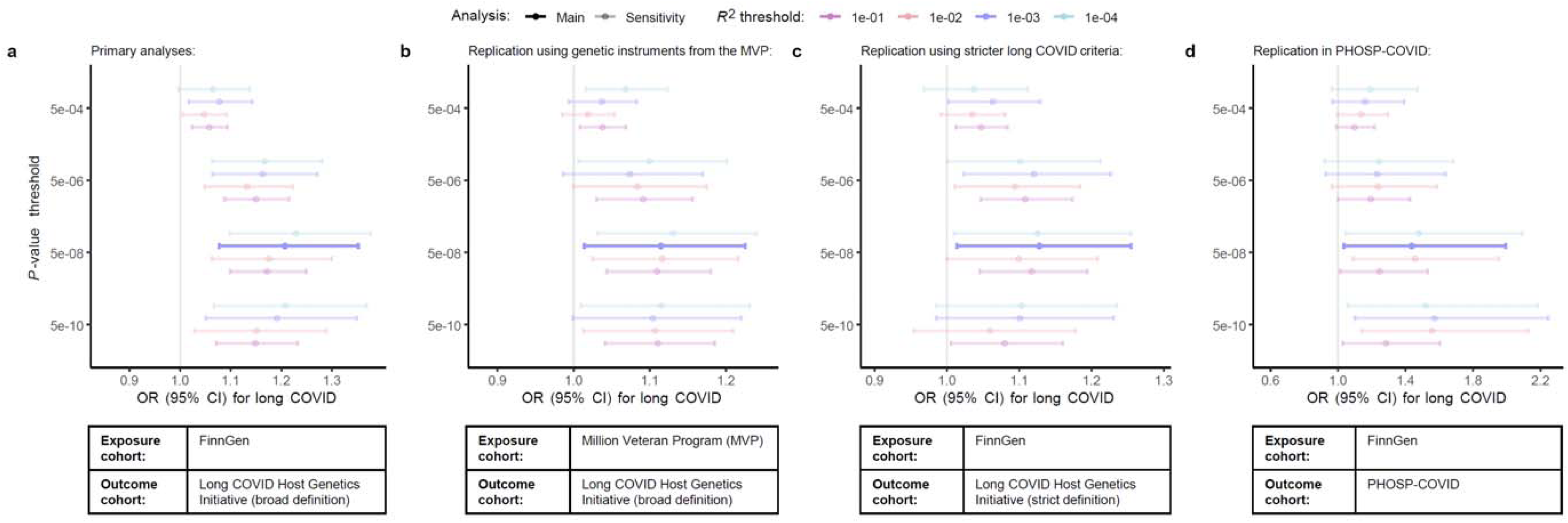
Associations of genetically predicted venous thromboembolism with long COVID in sensitivity and replication analyses. Sensitivity analyses tested the association of genetically predicted venous thromboembolism with long COVID using the inverse-variance weighted method with genetic instruments constructed using a range of *P*-value (*y*-axis) and linkage disequilibrium *R*^2^ (colors) thresholds to construct genetic instruments. The *P*-value threshold determines the minimal association strength for genetic variants included in each genetic instrument, whereas the linkage disequilibrium *R*^2^ threshold determines the maximal allowed correlation between genetic variants in each genetic instrument. **a**, In primary analyses, associations were calculated using genetic instruments for venous thromboembolism constructed in 412,181 participants from FinnGen^21^ and tested against long COVID ascertained in the Long COVID Host Genetics Initiative (“broad control definition”; *N*=997,600)^24^. Replication analyses used: **b**, genetic instruments from an external genotyped cohort with ascertainment of venous thromboembolism events (Million Veteran Program [MVP]; *N*=190,266)^27^; **c**, an alternative, stricter, control definition for long COVID in the Long COVID Host Genetics Initiative (*N*=40,910)^24^; and **d**, data from an external genotyped cohort with long COVID ascertainment (PHOSP-COVID; *N*=1,097)^28^. CI indicates confidence interval; OR, odds ratio.

Additionally, we tested for reverse causation (i.e., long COVID increasing venous thromboembolism risk) by applying Steiger filtering. While Steiger filtering can be used to identify variants explaining more variance in the outcome than in the exposure of interest^26^, we did not identify any such reverse causal variants. Correspondingly, MR analyses in the opposite direction (see **Methods**)—evaluating the effect of genetically predicted long COVID on venous thromboembolism risk—yielded null results (OR, 1.01 [95%CI, 0.94–1.09] per log-odds increase in genetically predicted long COVID risk; *P*=0.79), providing no evidence for reverse causation.

Replication analyses evaluated whether the observed associations were consistent across different exposure and outcome datasets (see **Methods**). First, we constructed an externally derived genetic instrument for venous thromboembolism using data from the Million Veteran Program^27^ and tested its association with long COVID in the Long COVID Host Genetics Initiative (**Supplementary Table 5**). Second, we tested the genetic association of venous thromboembolism with long COVID in a subcohort of the Long COVID Host Genetics Initiative that applied stricter criteria for controls, restricting to participants with proven SARS-CoV-2 infection^24^ (**Supplementary Table 6**). Third, we tested this association in the Post-Hospitalization COVID-19 (PHOSP-COVID) study, a prospective cohort study designed to investigate the medium- and long-term sequelae of hospitalized COVID-19^28,29^ (**Supplementary Table 7**). All three replication analyses demonstrated a significant positive association of genetically predicted venous thromboembolism risk with long COVID (**Fig. 3**). Additional sensitivity analyses, performed using the same replication datasets with varying linkage disequilibrium *R*^2^ and *P*-value thresholds for genetic instrument construction, yielded consistent results throughout (**Fig. 3**; **Supplementary Tables 5-7**).

### The thromboembolism-long COVID link does not reflect acute COVID-19 severity

Given that thrombotic events are frequently observed in individuals with COVID-19, correlating partly with disease severity^30,31^, and that severe COVID-19 is a thought to be a causal risk factor for long COVID^24^, we investigated whether acute COVID-19 severity might mediate the genetic association of venous thromboembolism with long COVID. Specifically, we tested the effects of genetically predicted venous thromboembolism risk across three categories of acute COVID-19 severity in the COVID-19 Host Genetics Initiative^16,32^: SARS- CoV-2 infection, hospitalized COVID-19, and critical COVID-19 (defined as hospitalization with respiratory support or COVID-19-related death). Our results showed that genetically predicted venous thromboembolism risk had effect estimates within the 95%CI for its association with long COVID for both hospitalized COVID-19 (OR, 1.09 [95%CI, 1.02-1.16] per log-odds increase in genetically predicted venous thromboembolism risk; *P*=1.6×10^-2^) and critical COVID-19 (OR, 1.08 [95%CI, 0.99-1.17] per log-odds; *P*=6.6×10^-2^), but not for SARS-CoV-2 infection alone (**Fig. 4a**). Only the association with hospitalized COVID-19 reached nominal significance.

**Figure 4.**
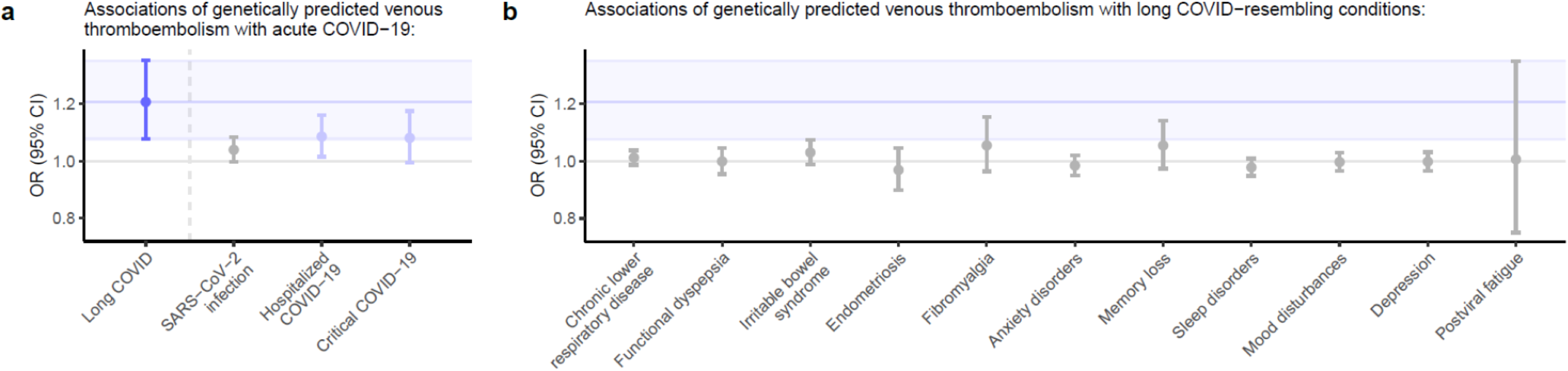
Analyses exploring the associations of genetically predicted venous thromboembolism with acute COVID-19 phenotypes and long COVID-resembling conditions. Analyses testing the associations of genetically predicted venous thromboembolism with **a**, long COVID and acute COVID-19 phenotypes and **b**, long COVID-resembling symptoms and conditions. For **a**, long COVID was ascertained in 997,700 participants from the Long COVID Host Genetics Initiative^24^ and the acute COVID-19 phenotypes were ascertained in up to 2,597,856 participants from the COVID-19 Host Genetics Initiative^32^. The genetic instruments for venous thromboembolism were constructed using data from 412,181 FinnGen participants^21^. For **b**, all long COVID-resembling conditions were ascertained in up to 412,181 participants from FinnGen^21^. The genetic instruments for venous thromboembolism were constructed using data from 190,266 Million Veteran Program^27^ participants to avoid sample overlap. For **a** and **b**, the *x*-axes highlight the different outcomes that were tested. All genetic instruments for the indicated conditions were constructed using genetic variants that were strongly associated with venous thromboembolism (*P*<5×10^-8^) and not in linkage disequilibrium with each other (linkage disequilibrium *R*^2^<0.001). All associations were calculated using the inverse-variance weighted Mendelian randomization method, visualized using odds ratios (ORs) and 95% confidence intervals (CIs). As shown in Fig. 2, genetically predicted venous thromboembolism was significantly associated with a higher risk of long COVID (depicted in dark blue; the blue band corresponds to the 95%CI of this association). Only “hospitalized COVID-19” and “critical COVID-19” (depicted in light blue) had associations that fell within the 95%CI of long COVID’s association.

Building on these findings, we performed multivariable MR^33^ to estimate the direct effects of genetic predisposition to venous thromboembolism on long COVID while adjusting for its impact on acute COVID-19 severity. Consistent with previous findings^24^, all acute COVID-19 phenotypes had significant genetic associations with long COVID in these multivariable MR models (**Supplementary Table 8**). However, genetically predicted venous thromboembolism was also significantly associated with long COVID across all models (**Supplementary Table 8**), suggesting that its effects on long COVID are not mediated by acute COVID-19 severity alone. These findings imply that a predisposition to thromboembolism contributes to the risk of developing long COVID through mechanisms that are not solely dependent on the severity of acute infection.

### Predisposition to thromboembolism does not promote symptoms shared with long COVID

To gain additional insights into the mechanisms driving the genetic association of venous thromboembolism with long COVID, we tested the associations of genetically predicted venous thromboembolism risk with specific conditions with symptom profiles shared with long COVID. Genetically predicted risk of venous thromboembolism was not associated with a higher risk of long COVID-resembling conditions or symptoms such as memory loss or mood disorders (**Fig. 4b**; **Supplementary Tables 9-10**). Although there was no significant association with postviral fatigue (OR, 1.13 [95%CI, 0.88-1.44] per log-odds increase in genetically predicted venous thromboembolism risk; *P*=0.35), confidence intervals were very wide, suggesting this association was potentially underpowered due to a small number of cases in FinnGen’s GWAS of postviral fatigue (*n*_cases_=195; *n*_controls_=382,198). Nevertheless, these findings suggests the genetic association of venous thromboembolism with long COVID represents a specific association, wherein thromboembolism plays a role in the pathophysiology of long COVID rather than the occurrence of its individual, non-specific symptoms.

### Exploration of potential molecular mediators

Finally, we aimed to identify potential molecular mediators of the thromboembolism- long COVID association by evaluating the effects of individual genes and their associated proteins on long COVID risk. First, we constructed “gene-specific genetic instruments” by identifying variants strongly associated with venous thromboembolism within or near different coagulation-related genes (see **Methods**). Gene-specific instruments consisting of one or multiple independent variants were available for a total of 40 genes (**Supplementary Tables 11-12**). Most genes’ instruments included only one genetic variant (*n*=32/40 [80.0%]). All mean *F*-statistics of the gene-specific instruments were greater than 10 (mean, 65.2; range, 12.9-377.4), indicating adequate instrument strength^10^. Among the different gene-specific instruments, *F2R*’s instrument (rs73131633) had the strongest effect on long COVID risk (**Fig. 5a**). Each unit increase in the log-odds of genetically predicted venous thromboembolism risk mediated through *F2R* was nominally associated with 4.50-fold odds of developing long COVID (95%CI, 1.12-18.2; *P*=3.5×10^-2^).

**Figure 5.**
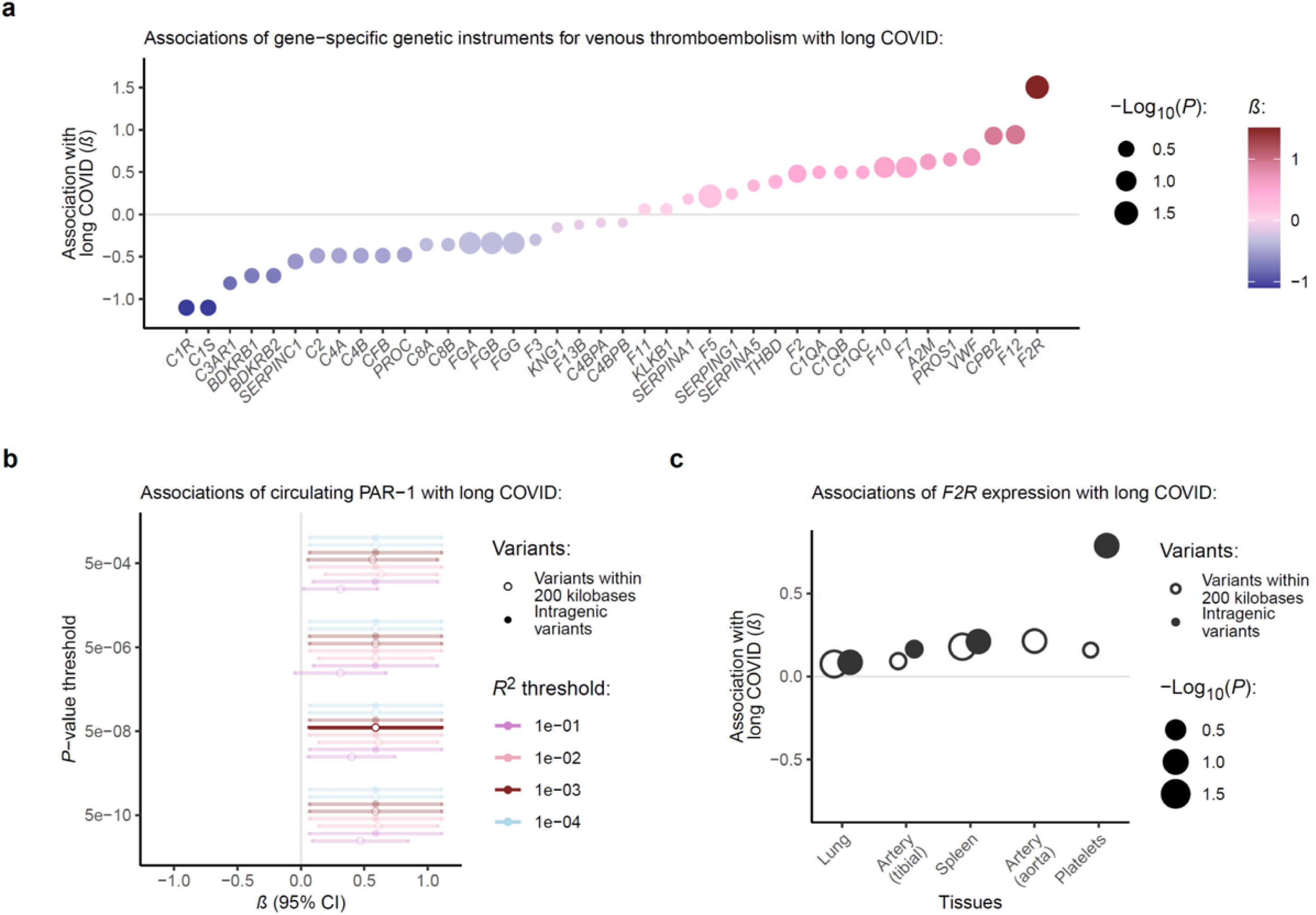
Identification of thromboembolism-related molecular mediators of long COVID. **a**, To identify coagulation-related genes driving the association of genetically predicted venous thromboembolism with long COVID, we tested the effects of gene-specific instruments for venous thromboembolism with long COVID. These gene-specific instruments were constructed using independent genetic variants associated with venous thromboembolism in FinnGen^21^ within 200 kilobases of the indicated gene. The *x*-axis depicts the different coagulation- and complement-related genes for which gene-specific instruments could be constructed. The *y*-axis and color gradient depict the effect size (*β*) of each association. The circle size depicts the negative log_10_-transformed *P*-value for each association. The greatest effect size was observed for *F2R*, which encodes the protease-activated receptor 1 (PAR-1) and was carried forward for further analyses. **b**, We tested the associations of genetically predicted circulating levels of PAR-1 with long COVID. Primary analyses tested a genetic instrument constructed using independent (*R*^2^<0.001) *cis-*variants (within 200 kilobases of *F2R*) associated wih circulating PAR-1 levels (*P*<5×10^-8^) in the UK Biobank Pharma Proteomics Project (UKB-PPP)^37^. Sensitivity analyses tested instruments constructed using a range of *P*-value (*y*-axis) and linkage disequilibrium *R*^2^ (different colors) thresholds as well as those constructed using only intragenic variants (different shapes). The *P*-value threshold determines the minimal association strength for genetic variants included in each genetic instrument, whereas the linkage disequilibrium *R*^2^ thresholds determines the maximal allowed correlation between genetic variants in each genetic instrument. **c**, Additional analyses tested the associations of genetically predicted expression levels of *F2R* in physiologically relevant tissues and blood cell types. Genetic instruments were constructed using independent genetic variants associated with expression levels of *F2R* in different tissues and platelets in GTEx^40^ and the GeneSTAR Research Study^41^, respectively. Analyses were performed using genetic instruments with intragenic variants or variants within 200 kilobases of *F2R* (different shapes). The *y*-axis and color gradient depict the effect size (*β*) of each association. The circle size depicts the negative log_10_-transformed *P*-value of each association. For **a**, **b**, and **c**, all genetic instruments were tested in the Long COVID Host Genetics Initiative^24^ using the inverse-variance weighted method.

The *F2R* gene encodes the protease-activated receptor 1 (PAR-1; i.e., the thrombin receptor). PAR-1 is a G-protein-coupled receptor that is activated predominantly by thrombin and drives thrombosis and inflammation through involvement in platelet aggregation, endothelial activation, and leukocyte recruitment^34^. To verify its association with long COVID, we tested the effects of genetically predicted circulating levels of PAR-1 on long COVID risk. Genetically predicted circulating levels of PAR-1 were instrumented using a single intronic *F2R* variant (rs168753; in moderate linkage disequilibrium [*R*^2^=0.36] with rs73131633) associated with modified PAR-1 expression on platelets leading to altered procoagulant activity^35,36^; this variant was also the lead *F2R* signal for circulating PAR-1 levels in the UK Biobank Pharma Proteomics Project^37^. MR analyses using this instrument revealed that each unit increase in genetically predicted PAR-1 levels was associated with 1.80-fold odds of developing long COVID (95%CI, 1.07-3.04; *P*=2.7×10^-2^). This association was consistent across genetic instrument construction parameters (**Fig. 5b**; **Supplementary Table 13**).

Colocalization analyses supported the hypothesis of a causal variant for trait 1 (i.e., circulating PAR-1 levels) but not trait 2 (i.e., long COVID) as the most likely (75.8%), suggesting that statistical power was insufficient to discriminate between whether the genetic association was due to shared causal variants or variants in linkage disequilibrium (i.e., horizontal pleiotropy)^38,39^. The probability for shared causal variants, conditional on the presence of a causal variant for both traits, was 60.9%.

We also evaluated the association between genetically predicted *F2R* expression and long COVID risk. Using data from the Genotype-Tissue Expression (GTEx) project^40^, we identified the tissues with the highest levels of *F2R* expression and constructed genetic instruments for those most relevant to long COVID pathophysiology. As GTEx lacks data on blood platelets—an important site of *F2R* expression^34^—we supplemented our analysis with genetic instruments for platelets from the GeneSTAR Research Study^41^. Each tissue-specific instrument was constructed using the variant most strongly associated with *F2R* expression within a 200-kilobase *cis*-region of the gene. As *F2R*’s *cis*-region partially overlaps with those of other genes (e.g., *F2RL1* and *F2RL2*)^42^, we also performed analyses using only intragenic variants. Our analyses revealed that genetically predicted *F2R* expression had positive effect estimates on long COVID risk across all tissues, with nominal significance for *F2R* expression in platelets, lung, and spleen tissue (**Fig. 5c**; **Supplementary Table 14**). The strongest effect was observed in platelets, indicating that genetic variants increasing *F2R* expression in platelets are linked to higher long COVID risk, further supporting a mechanistic link between thromboembolism and long COVID.

## Discussion

Using genetic data from multiple cohorts, we tested the association of genetically predicted risk of venous thromboembolism with long COVID. Our primary analysis demonstrated that a higher genetically predicted risk of venous thromboembolism was significantly associated with an increased risk of developing long COVID. This association was robust across various sensitivity analyses, replicated using data from external cohorts, and was independent of acute COVID-19 severity. Additional analyses revealed no genetic associations of venous thromboembolism with any long COVID-resembling conditions (e.g., memory loss or mood disorders), suggesting a specific role for thromboembolism-related pathways in long COVID. Furthermore, gene-focused analyses identified PAR-1—a thrombin receptor strongly involved in platelet and coagulation activation—as a potential contributor to its pathophysiology. Collectively, our findings provide new mechanistic insights and identify a potential therapeutic target for long COVID.

These results have several implications. First, our findings suggest that thromboembolic pathways are causally involved in the development of long COVID. This extends earlier observational research linking thromboinflammatory biomarkers to long COVID^6,7,43^. For instance, prior analyses have shown that blood biomarkers of coagulation and inflammation (i.e., fibrinogen and D-dimer relative to C-reactive protein) can help predict cognitive defects in patients hospitalized for COVID-19 up to 12 months after infection^6^.

Similarly, a recent prospective study of 113 COVID-19 patients found that individuals who developed long COVID exhibited changes to blood proteins indicative of persistent thromboinflammation at 6 months post-infection, compared to those who did not develop long COVID^7^. However, these studies relied predominantly on traditional observational approaches to identify molecular risk factors for long COVID. While such approaches are often well-suited to identify biomarkers for the detection and prediction of diseases, they cannot always discern true causal associations from those influenced by confounding or reverse causation^10^. MR is an approach that can help deal with these challenges by using genetic variants as instruments for specific exposures. Because an individual’s genetic makeup is fixed at conception, MR analyses are less directly susceptible to external influences and reverse causation than those performed using more traditional methods^10^. Consequently, this study provides human genetic evidence supporting a causal association of thromboembolism with long COVID.

Second, while thrombotic events are common in individuals with acute COVID-19 and correlate partially with disease severity^30,31^, our findings indicate that thromboembolic pathways may contribute to long COVID independently of acute disease severity. During the early stages of the pandemic, one in four COVID-19 patients admitted to the intensive care unit experienced venous thromboembolism despite standard dose thromboprophylaxis^44^.

Mechanistic research suggests that this heightened risk of venous thromboembolism partially reflects a systemic COVID-19-associated coagulopathy involving immune response dysregulation, endothelial cell dysfunction, and hypercoagulability^45^. For instance, early work demonstrated that SARS-CoV-2 infection is associated with platelet hyperreactivity (e.g., in response to PAR-1 stimulation)^46^ potentially contributing to the formation of microthrombi across organs (e.g., the lungs and brain)^45,47–49^. Of note, epidemiological research indicates that individuals with a genetic predisposition to venous thromboembolism are more likely to experience acute thrombotic complications due to COVID-19^50^, which aligns with our observation that genetic predisposition to venous thromboembolism is associated with an increased risk of hospitalized COVID-19. However, we also found that the genetic association of venous thromboembolism with long COVID was not mediated by acute disease severity. This finding is consistent with *in vitro* evidence showing that the SARS-CoV-2 spike protein binds human fibrin, promoting persistent thromboinflammation and neuropathology independent of active infection^51^. Together with recent reports of circulating SARS-CoV-2 spike in individuals with long COVID^52^, our findings support the hypothesis that SARS-CoV-2 creates a substrate for sustained thromboinflammation post-infection, potentially leading to chronic symptoms from multiple organ systems.

Third, gene-focused analyses identified PAR-1 as a potential contributor to the pathogenesis of long COVID. PAR-1 is encoded by the *F2R* gene and represents the prototype of a small family of G-protein-coupled protease-activated receptors (PARs) involved in thrombin signaling^34,53^. Thrombin cleaves the amino-terminal extracellular domain of PAR-1, resulting in the formation of a new amino terminus that binds intramolecularly to the body of the receptor and leads to platelet activation^53^. While PAR-1 predominantly functions as a receptor and not conventionally as a secreted protein in the circulation, we found that higher genetically predicted plasma levels of this protein (e.g., through receptor shedding or in extracellular vesicles) were associated with a higher risk of long COVID. As PAR-1 is considered the main mediator of thrombin’s effects on platelet aggregation, it has gained interest as a potential drug target for the prevention of thrombotic events^54,55^. Large trials found that inhibition of PAR-1 with vorapaxar (i.e., a competitive PAR-1 antagonist) reduced the risk of cardiovascular death or ischemic events in selected populations^54^; however, it also increased the risk of major bleeding including intracranial hemorrhage^54,55^. In addition to mediating thrombin-induced platelet aggregation, PAR-1 also promotes coagulation-driven inflammation. For instance, in an experimental mouse model of sepsis, genetic and pharmacological inhibition of PAR-1 led to reduced inflammation and coagulation in comparison with control treatment, as did pharmacological inhibition of thrombin^56^.

Consistent with these findings, data from recent mechanistic work suggest that virus-exposed endothelium promotes thromboinflammation through a positive feedback loop sustained and enhanced by thrombin and PAR-1, implicating PAR-1 in virus-induced inflammation and thrombosis^57^. Together with the results from the present study, these findings support the idea that PAR-1 is a mediator of virus-induced thromboinflammation, potentially contributing to chronic symptoms associated with SARS-CoV-2 infection. Future animal experiments and clinical trials are needed to evaluate the effectiveness of drugs targeting PAR-1 or other thromboinflammatory mediators to prevent and/or treat long COVID.

Although this study benefitted from multiple large-scale and well-profiled datasets, as well as a robust framework for causal inference using human genetics, certain limitations must be considered when interpreting the study findings. First, while MR analyses can be used to infer causality in exposure-outcome pairs, causal inference relies on the justification of the underlying MR assumptions^10^. However, the present study used a robust framework with multiple sensitivity and replication analyses and assessed the key MR assumptions where possible (see **Methods**), minimizing the chance of systematically violating the MR assumptions. Second, we only tested genetic instruments constructed using data from European-ancestry cohorts, limiting generalizability to other ancestries. Third, certain phenotypes such as (post-viral) post-exertional malaise did not have GWAS data available and could therefore not be evaluated in relation to genetically predicted venous thromboembolism risk. Fourth, the use of partially overlapping cohorts can lead to bias towards the observational exposure-outcome association; however, given the limited number of long COVID cases in FinnGen (*n*_cases_=263), the binary nature of the outcome phenotype (i.e., long COVID), and the strong genetic instrument for venous thromboembolism (mean *F*- statistic, 114.5), any bias due to sample overlap was expected to be minimal^58^. Furthermore, replication analyses with non-overlapping exposure and outcome datasets (i.e., the Million Veteran Program and PHOSP-COVID cohorts) yielded highly consistent results, suggesting sample overlap did not strongly affect our findings. Fifth, although the gene-specific association of venous thromboembolism with long COVID in *F2R* had a large effect size, this association did not reach Bonferroni-significance due to limited power. Similarly, colocalization analyses to support the association of circulating PAR-1 and long COVID had limited power. Nevertheless, the probability for shared causal variants, conditional on the presence of a causal variant for long COVID, was 61.0%, which means that the probability for shared causal variants was greater than that of distinct causal variants and suggests no confounding due to linkage disequilibrium. Finally, while the gene-focused analyses from the present study used a robust *cis*-MR framework facilitating adherence to the core assumptions of MR^59–61^, the identification of PAR-1 as a potential therapeutic target should be validated in prospective intervention trials.

In conclusion, this study found a strong and robust genetic association of venous thromboembolism with long COVID, pointing to thromboembolic pathways as potential drivers of post-acute sequelae following SARS-CoV-2 infection. The thrombin receptor PAR- 1 was identified as a putative molecular contributor to long COVID pathogenesis. These findings offer human genetic evidence for a causal link between thromboembolism and long COVID and support clinical research into anticoagulant and antiplatelet therapies as strategies for its prevention and treatment.

## Methods

### MR and instrumental variable assumptions

MR analyses can be used to infer causality in certain exposure-outcome relationships, given that these analyses adhere to three key instrumental variable assumptions^10^ (**Fig. 1**). First, the tested genetic instrument must comprise genetic variants that are associated with the exposure of interest (i.e., relevance assumption). Second, there can be no confounding pathway between the genetic variants comprising the tested genetic instrument and the outcome (i.e., independence assumption). Third, there can be no horizontal pleiotropy in the association of the tested genetic instrument with the outcome, meaning that the tested genetic instrument can only affect the outcome through its effects on the exposure of interest (i.e., exclusion restriction assumption).

The relevance assumption was tested by estimating the *F*-statistics (indicating instrument strength) for the variants included in the genetic instrument for venous thromboembolism^10^. The independence and exclusion restriction assumptions were evaluated by testing the associations of the genetic instrument for venous thromboembolism with a range of long COVID-resembling conditions^10^. We also performed replication analyses using a wide range of genetic instruments and pleiotropy-robust methods (e.g., MR-Egger^25^) to further evaluate the robustness of the observed associations against the aforementioned MR assumptions.

### GWAS of venous thromboembolism

Genetic association data for venous thromboembolism were obtained from FinnGen^21^. The FinnGen study is a large-scale genomics initiative that has analyzed over 500,000 Finnish biobank samples and correlated genetic variation with health data to understand disease mechanisms and predispositions. The present study used data from FinnGen data release 10, which includes 412,181 participants with genetic and phenotype data. The Coordinating Ethics Committee of the Hospital District of Helsinki and Uusimaa (HUS) approved the FinnGen study protocol, and all participants provided informed consent^21^.

The genetic instrument for venous thromboembolism (I9_VTE) was constructed using a GWAS in 21,021 venous thromboembolism cases and 391,160 controls^23^. Venous thromboembolic events were defined using diagnostic codes from hospital discharge and death cause registers. In brief, qualifying events included diagnoses of pulmonary embolism and deep vein thrombosis. The exact codes used to construct the venous thromboembolism phenotype are listed in **Supplementary Table 15**. Participants were genotyped using different Illumina (Illumina Inc., San Diego, CA, USA) and Affymetrix arrays (Thermo Fisher Scientific, Santa Clara, CA, USA)^62^. Samples with ambiguous gender, high genotype missingness (>5%), excess heterozygosity (more than 4 standard deviations from the mean) and non-Finnish ancestry were excluded^62^. Samples were imputed to the 1000 Genomes Project reference panel (phase 3)^63^ prior to association analyses adjusted for age, sex, ten principal components of ancestry, FinnGen chip version 1 or 2, and legacy genotyping batch as covariates^62^. Variants with high missingness (>2%) and Hardy-Weinberg equilibrium *P*<1×10^-6^ were excluded^62^. Variants with minor allele frequency <1% were also excluded.

### GWAS of long COVID

Genetic association data for long COVID were obtained from the Long COVID Host Genetics Initiative^24^. Data from a total of 3,018 long COVID cases and 994,582 controls from 12 individual cohorts were included for primary analyses (**Supplementary Table 1**).

Consistent with the genetic ancestry of the exposure cohort, more than 99% of participants had European ancestry. The maximal overlap in effective sample size (calculated as [4 × *n*_case_ × *n*_control_] / [*n*_case_ + *n*_control_]^24^) between the Long COVID Host Genetics Initiative and FinnGen^21^ cohorts was 10.3%. Given the limited number of long COVID cases in FinnGen (*n*_cases_=263), the binary nature of the outcome phenotype (i.e., long COVID), and the strong genetic instrument for venous thromboembolism (mean *F*-statistic, 114.5), any bias due to sample overlap is expected to be minimal^58^. Furthermore, we performed replication analyses with different exposure and outcome datasets (i.e., the Million Veteran Program and PHOSP-COVID) without any sample overlap (see **Replication analyses**). Given that these datasets were smaller—and less well-powered—than the FinnGen and Long COVID Host Genetics Initiative datasets, we chose to use the latter set of datasets for primary analyses and the other for replication analyses. Long COVID Host Genetics Initiative participants provided informed consent for participation in their respective studies, with recruitment and ethics following study-specific protocols approved by their respective institutional review boards^24^.

Study participants were classified as long COVID cases if they had an earlier test-verified SARS-CoV-2 infection and met long COVID criteria at least three months since SARS-CoV-2 infection or COVID-19 onset^24^. Aligning with the World Health Organization guidelines^2^, individuals meeting long COVID criteria were defined as those who (1) reported presence of COVID-19 symptoms that could not be explained by alternative diagnoses; (2) reported ongoing significant impact on day-to-day activities; or (3) had any diagnosis codes of long COVID in their electronic health records. Primary analyses used population controls (i.e., all non-cases) as controls; these individuals were defined as those who were not identified as having long COVID using the above-mentioned criteria. As such, data from a total of 3,018 long COVID cases and 994,582 controls from 12 cohorts were included for primary analyses. All cohort-specific criteria to ascertain long COVID are shown in **Supplementary Table 1**. Each cohort study applied their own methods for genotyping, genotype and sample quality control, imputation, and association analyses (**Supplementary Table 1**), according to a central analysis plan^24^. Each cohort’s GWAS was minimally adjusted for age, age^2^, sex, age × sex, and the first 10 genetic principal components. Prior to analysis, all variants with imputation INFO score <0.6 were excluded. Meta-analysis was performed using a fixed-effects inverse-variance weighted method.

Primary analyses used population controls to increase statistical power and limit selective inclusion of certain groups within the population, which can introduce collider bias in MR studies. Population controls were defined as those who were not identified as having long COVID using the above-mentioned criteria^24^. Replication analyses used a stricter control definition (i.e., all individuals with a history of SARS-CoV-2 infection but without long COVID criteria [*n*_cases_=2,975; *n*_controls_=37,935]). Additional details on the long COVID case and control phenotypes, including diagnostic codes and study-specific criteria, can be found in the **Supplementary Table 1** and in the main Long COVID Host Genetics Initiative paper^24^.

### MR analyses

In our primary analysis, we tested the association of genetically predicted venous thromboembolism risk with long COVID. To do so, we constructed a genetic instrument (i.e., a statistical entity comprising one or more genetic variants) for venous thromboembolism in FinnGen^21^. All genetic instrument construction steps were performed using genetic variants that were present in both the exposure and outcome GWAS datasets. To ensure that our genetic instrument was strongly associated with venous thromboembolism, we only included variants that were associated with venous thromboembolism at genome-wide significance (*P*<5×10^-8^). In addition, to limit bias caused by genetic variants in linkage disequilibrium, we only retained independent variants in our genetic instrument (*R*^2^<0.001; clumped within regions of 10 megabases using PLINK [version 1.90]^64^). The linkage disequilibrium reference panel for clumping was derived from the European panel of phase 3 of the 1000 Genomes Project^63^.

Primary analyses used the inverse-variance weighted method with multiplicative random effects^12^ to estimate the effects of genetically predicted venous thromboembolism risk on long COVID. In downstream analyses, unless specified otherwise, all MR analyses testing the effects of a genetic instrument with more than three variants used this method. MR analyses testing the effects of a genetic instrument with two or three variants used the inverse-variance weighted method with fixed effects^12^; those testing the effects of a genetic instrument with only one variant used the Wald ratio estimator^65^.

All associations were presented as odds ratios (ORs) and 95% confidence intervals (CIs) per log-odds increase in genetically predicted risk for the exposure condition, along with their corresponding *P*-values. Two-sided *P*<0.05 indicated statistical significance in primary analyses (i.e., the inverse-variance weighted estimate of genetically predicted venous thromboembolism risk on long COVID). All MR analyses were performed using the *TwoSampleMR*^66^ (version 0.5.7) and *MendelianRandomization*^67^ (version 0.7.0) packages in R (version 4.1.0) were used for analyses.

### Genetic instruments for non-venous thromboembolism conditions

To contextualize the genetic association of venous thromboembolism with long COVID (i.e., the association observed in our primary analysis), we also tested the genetic associations of different other conditions with long COVID. These conditions were chosen from FinnGen^21^ to represent a variety of organ systems and pathophysiological pathways with well-powered GWAS and included: atrial fibrillation (FinnGen phenotypee code: I9_AF), asthma (J10_ASTHMA_EXMORE), chronic kidney disease (N14_CHRONKIDNEYDIS), chronic lower respiratory disease (J10_LOWCHRON), coronary artery disease (I9_REVASC), dementia (F5_DEMENTIA), diabetes mellitus (T2D), hypertension (I9_HYPTENS), migraine (MIGRAINE_TRIPTAN), multiple sclerosis (G6_MS), osteoarthritis (M13_ARTHROSIS), and rheumatoid arthritis (M13_RHEUMA) (**Supplementary Table 15**).

Consistent with the primary analysis, all genetic instruments for these conditions were constructed using GWAS in FinnGen (release 10)^21^. We also tested the associations of genetically predicted risk for deep vein thrombosis (I9_PHLETHROMBDVTLOW) and pulmonary embolism (I9_PULMEMB) with long COVID to corroborate the genetic association of venous thromboembolism with long COVID. All tested phenotypes were defined using nationwide registries that were linked to the biological samples and harmonized across diagnosis-, procedure-, and drug-specific codes. The exact codes used to construct the different phenotypes are listed in **Supplementary Table 15**. The GWAS for these phenotypes (including genotyping, variant- and sample-wise quality control steps, and association analyses) were identical to those used for the GWAS of venous thromboembolism in FinnGen (see **GWAS of venous thromboembolism**). All GWAS were well-powered with an adequate number of cases per condition (*n*>2,000) and multiple independent (*R*^2^<0.001) variants reaching genome-wide significance (*P*<5×10^-8^).

All genetic instruments for these conditions were constructed using a *P*-value threshold of 5×10^-8^ and a linkage disequilibrium *R*^2^ threshold of 0.001. The associations of these genetic instruments with long COVID were tested using the inverse-variance weighted method. As the main aim of these analyses was to compare the association observed for venous thromboembolism (i.e., our primary analysis) with the associations observed for other, unrelated conditions, statistical significance for these analyses was set at the same threshold (i.e., two-sided *P*<0.05). In addition, to compare effect estimates, we also evaluated which genetic instruments had effect estimates within the 95%CI of the venous thromboembolism-long COVID association.

### Sensitivity analyses

We performed sensitivity analyses using different MR approaches and genetic instrument selection parameters to test the robustness of the association of genetically predicted venous thromboembolism risk with long COVID. First, to evaluate the possibility of horizontal pleiotropy (i.e., effects of a genetic instrument on the target outcome beyond its effects on the exposure of interest) affecting the observed associations, we performed sensitivity analyses using MR methods that allow for horizontal pleiotropy (including MR-Egger, the weighted median estimator, and the weighted mode estimator). MR-Egger is an MR method used to test and correct for horizontal pleiotropy within a genetic instrument at the cost of lower precision and statistical power^25^. The weighted median estimator is an alternative MR method with preserved statistical power that allows for horizontal pleiotropy as long as the majority of the genetic instrument’s weight consists of valid instrumental variables (i.e., genetic variants)^68^. The weighted mode-based estimator allows for horizontal pleiotropy as long as a plurality of the genetic instrument’s weight consists of valid instrumental variables^69^. Second, to explore the potential effects of residual correlation between the genetic variants in our genetic instrument, we conducted sensitivity analyses with genetic instruments constructed using various linkage disequilibrium *R*^2^ thresholds (*R*^2^<0.0001/*R*^2^<0.001/*R*^2^<0.01/*R*^2^<0.1). Third, we also conducted additional sensitivity analyses with genetic instruments constructed using various *P*-value thresholds (*P*<5×10^-4^/*P*<5×10^-6^/*P*<5×10^-8^/*P*<5×10^-10^). These sensitivity analyses were performed in line with previous reports^59,60,70^. Although there is no consensus on the exact values to use for these sensitivity analyses, we chose the aforementioned *R*^2^ and *P*-value thresholds to cover a reasonable range of parameters at fixed intervals. As MR analyses using multiple genetic variants from across the genome conventionally use a genome-wide significance threshold (*P*<5×10^-8^) and a relatively stringent *R*^2^ threshold (*R*^2^<0.001), the different *R*^2^ and *P*-value parameters for sensitivity analyses were centered around these values. Fourth, we performed Steiger filtering, which removes genetic variants that explain more variation in the outcome than in the exposure of interest^26^, to evaluate the possibility of reverse causation affecting the observed results. Fifth, we performed MR analyses in the opposite direction (i.e., with long COVID as the exposure and venous thromboembolism as the outcome) to further explore the possibility of reverse causation. Genetic instruments were constructed using independent (*R*^2^<0.001) genome-wide significant (*P*<5×10^-8^) variants associated with long COVID in the Long COVID Host Genetics Initiative (**Supplementary Table 16**), using the same GWAS data as those used for the outcome in primary analyses. These reverse MR analyses used the inverse-variance weighted method. Because the genetic instrument for long COVID constructed using a *P*-value threshold of 5.0×10^-8^ (mean *F*-statistic, 35.2) only included two variants, we also performed an additional Mendelian randomization analysis using a *P*-value threshold of 5.0×10^-6^ for instrument construction (mean *F*-statistic, 23.8), yielding similar results (OR, 0.99 [95%CI, 0.95-1.03] per log-odds increase in long COVID risk; *P*=0.52).

Sensitivity analyses were considered robust if (1) MR-Egger suggested no horizontal pleiotropy (*P*≥0.05 for the intercept test or *P*<0.05 for the intercept test with *P*<0.05 for the causal test) and all pleiotropy-robust methods (i.e., the MR-Egger, weighted median, and weighted mode-based estimators) had a consistent direction of effect; (2) MR estimates were directionally consistent across all linkage disequilibrium *R*^2^ thresholds for instrument construction; (3) MR estimates were directionally consistent across all *P*-value thresholds for instrument construction; (4) MR analyses with Steiger filtering yielded directionally consistent results (or Steiger filtering did not identify any “reverse causal” variants); and (5) there was no significant positive association of genetically predicted long COVID risk on venous thromboembolism.

### Replication analyses

Replication analyses evaluated whether the observed associations were consistent across different exposure and outcome datasets. First, we constructed a genetic instrument for venous thromboembolism using a data source external to the FinnGen^21^ dataset (i.e., the data source used to construct the genetic instrument used for our primary analyses). To this end, we used data from 8,929 venous thromboembolism cases and 181,337 controls with European ancestry from the Million Veteran Program^27^. In brief, the Million Veteran Program recruited individuals aged 19 to >100 years from more than 50 Veterans Affairs Medical Centers across the USA since 2011^71^. Participants with at least two qualifying diagnostic codes for deep vein thrombosis or pulmonary embolism in their electronic health record were classified as cases^27^. Participants were genotyped using a customized Affymetrix Axiom array (Thermo Fisher Scientific, Santa Clara, CA, USA). Samples with excess heterozygosity, excess missingness (>2.5%), or discordance between genetically inferred sex and phenotypic gender were excluded^27^. Samples were imputed to the 1000 Genomes Project reference panel (phase 3)^63^ prior to association analyses adjusted for age, sex, and five principal components of ancestry^27^. Variants with ancestry-specific Hardy-Weinberg equilibrium *P*<1×10^-20^, posterior call probability <0.9, imputation INFO score <0.3, or call rate <97.5% were excluded, as were variants deviating >10% from their expected allele frequency based on reference data from the 1000 Genomes Project^63^. Variants with minor allele frequency <1% were also excluded. The genetic instrument for venous thromboembolism used for replication was constructed using the same approach as for the instrument used in our primary analysis (e.g., using a *P*-value threshold of 5×10^-8^ and a linkage disequilibrium *R*^2^ threshold of 0.001). The Million Veteran Program received ethical and study protocol approval by the Veterans Affairs Central Institutional Review Board, and informed consent was obtained from all participants^27^.

Second, we evaluated whether the observed associations persisted when we used stricter criteria for controls in the Long COVID Host Genetics Initiative GWAS^24^. Unlike our primary analyses, which used population controls (i.e., all study participants without long COVID criteria; *n*_controls_=994,582), these replication analyses only included proven controls (i.e., all study participants with proven SARS-CoV-2 infection but without long COVID criteria; *n*_controls_=37,935).

Third, we tested the association of genetically predicted venous thromboembolism risk (i.e., the FinnGen instrument) with long COVID in the Post-Hospitalization COVID-19 (PHOSP-COVID) study, a prospective cohort study designed to investigate the medium- and long-term sequelae of hospitalized COVID-19^28,29^. PHOSP-COVID recruited adults who were discharged from one of 53 hospitals across the United Kingdom between March 5 and November 30, 2020, after confirmed or clinician-diagnosed COVID-19^28,29^. A total of 1,097 participants with genetic data and information on long COVID status (*n*_cases_=697; *n*_controls_=400) were included in this analysis. Long COVID status was primarily based on the question “Do you feel fully recovered from COVID-19?” during the a visit occurring at least 3 months after COVID-19 diagnosis. Participants who answered “no” were classified as long COVID cases. For participants who responded with "not sure" or who had a missing response, long COVID status was determined by their answers to the EuroQol five-dimension five-level (EQ-5D-5L) questionnaire. At their first visit (i.e., at least 3 months after COVID-19 diagnosis), participants completed the EQ-5D-5L questionnaire for their health state at that moment and retrospectively for their health state before COVID-19 diagnosis. Patients were categorized as having long COVID if the difference between the current and pre-COVID-19 EQ-5D-5L scores was ≤-0.1. Samples were genotyped using the Illumina Global Screening Array v3.0 (Illumina Inc., San Diego, CA, USA). Samples with call rates <95%, discordance between genetically imputed and recorded sex, excess relatedness (third degree of kinship), or excess heterozygosity (more than 4 standard deviations from the mean) were excluded, as were duplicates. Genetic variants with Hardy-Weinberg equilibrium *P*<1×10^-6^, call rates <99%, or minor allele frequencies <1% were also excluded. Genetic association analyses for long COVID were adjusted for age, age^2^, sex, age × sex, and ten principal components of genetic ancestry. The PHOSP-COVID study was approved by the Leeds West Research Ethics Committee, and all participants provided informed consent^28,29^.

Two-sided *P*<0.05 indicated statistical significance for all replication analyses. In addition to the three main replication analyses, which used the same methods as those used for our primary analyses (i.e., genetic instruments with *P*<5×10^-8^ and *R*^2^<0.001 tested using the inverse-variance weighted method), we also performed additional analyses using different *P*-value (*P*<5×10^-4^/*P*<5×10^-6^/*P*<5×10^-8^/*P*<5×10^-10^) and linkage disequilibrium *R*^2^ (*R*^2^<0.0001/*R*^2^<0.001/*R*^2^<0.01/*R*^2^<0.1) thresholds to construct genetic instruments. Analyses were considered robust if they were directionally consistent across all genetic instrument construction parameters.

### MR analyses of acute COVID-19 phenotypes

We tested the associations of genetically predicted venous thromboembolism risk with acute COVID-19 in the COVID-19 Host Genetics Initiative (release 7)^16,32^. The COVID- 19 Host Genetics Initiative includes GWAS data for three acute COVID-19 categories based on disease severity: SARS-CoV-2 infection (defined as any SARS-CoV-2 infection with or without symptoms of any severity); hospitalized COVID-19 (defined as hospitalization due to symptoms associated with SARS-CoV-2 infection); and critical COVID-19 (defined as hospitalization with respiratory support or death due to symptoms associated with SARS-CoV-2 infection)^16,32^. All three acute COVID-19 severity categories were tested as separate outcomes. We only used genetic data from individuals with European ancestry to match the ancestry of the outcome cohort (i.e., the COVID-19 Host Genetics Initiative^16,32^) with that of the exposure cohort (i.e., FinnGen^21^). All protocols for studies included in the COVID-19 Host Genetics Initiative followed local ethics recommendations, and informed consent was obtained when required^16,32^.

The different GWAS meta-analyses—excluding the 23andMe samples—included 122,616 cases and 2,475,240 controls for SARS-CoV-2 infection; 32,519 cases and 2,062,805 controls for hospitalized COVID-19; and 13,769 cases and 1,072,442 controls for critical COVID-19^16,32^. Each contributing cohort genotyped the samples, performed variant-wise and sample-wise quality control, and conducted association analyses independently following a central analysis plan. The different cohort-specific GWAS were filtered for variants with imputation INFO scores >0.6 and meta-analyzed using the inverse-variance weighted method. Variants with minor allele frequency <1% were excluded. Consistent with our primary analysis, we tested the associations of our main genetic instrument for venous thromboembolism with the three aforementioned acute COVID-19 phenotypes using the inverse-variance weighted MR method.

We performed multivariable MR^33^ analyses to estimate the direct effects of genetic predisposition to venous thromboembolism on long COVID while adjusting for its impact on acute COVID-19. Multivariable MR is an extension of conventional (i.e., univariable) MR that allows for multiple exposures to be modelled at once^33^. We performed three sets of multivariable MR analyses; in each analysis, venous thromboembolism was modelled as an exposure together with one acute COVID-19 phenotype (i.e., SARS-CoV-2 infection, hospitalized COVID-19, and critical COVID-19) while long COVID was considered the outcome. Genetic instruments were constructed using independent variants (*R*^2^<0.001) reaching genome-wide significance (*P*<5×10^-8^) in either the GWAS of venous thromboembolism or the GWAS of the acute COVID-19 phenotype under study. The independent genetic effects of each exposure on long COVID were estimated using the inverse-variance weighted method for multivariable MR studies, implemented using the *mr_mvivw()* function from the *MendelianRandomization*^67^ package (version 0.7.0) in R.

As the main aim of the conventional (i.e., univariable) MR analyses was to compare the genetic effects of venous thromboembolism on long COVID (i.e., our primary analysis) with those on acute COVID-19 phenotypes, statistical significance for these analyses was set at the same threshold (i.e., two-sided *P*<0.05). We also evaluated which associations had effect estimates within the 95%CI of the venous thromboembolism-long COVID association. Results from multivariable MR analyses were considered robust if venous thromboembolism had a significant independent association (i.e., two-sided *P*<0.05) with long COVID across models.

### MR analyses of long COVID-resembling conditions

We tested the genetic associations of venous thromboembolism with a range of “long COVID-resembling” phenotypes. These long COVID-resembling phenotypes included various conditions sharing similar symptoms—yet likely distinct mechanisms—to those observed in long COVID^3^. The phenotypes were chosen based on prior evidence suggesting overlapping symptoms with long COVID^3^ and were tested if they had GWAS data available in FinnGen (release 10)^21^.

All phenotypes were defined using nationwide registries that were linked to the biological samples and harmonized across diagnosis-, procedure-, and drug-specific codes^21^. The tested phenotypes included: chronic lower respiratory disease (FinnGen phenotype code: J10_LOWCHRON), functional dyspepsia (K11_FUNCDYSP), irritable bowel syndrome (K11_IBS), endometriosis (N14_ENDOMETRIOSIS), fibromyalgia (M13_FIBROMYALGIA), anxiety disorders (F5_ALLANXIOUS), memory loss (MEMLOSS), sleep disorders (SLEEP), mood disturbances (F5_MOOD), depression (F5_DEPRESSIO), and postviral fatigue (G6_POSTVIRFAT). Additional information on the tested long COVID-resembling phenotypes can be found in **Supplementary Table 10**. The GWAS for these phenotypes (including genotyping, variant-wise and sample-wise quality control, and association analyses) were identical to those used for the GWAS of venous thromboembolism in FinnGen (see **GWAS of venous thromboembolism**).

To avoid sample overlap, we used the genetic instrument from the Million Veteran Program^27^ (see **Replication analyses**) to test the effects of genetically predicted venous thromboembolism risk on long COVID-resembling phenotypes. These analyses used the inverse-variance weighted MR method. Statistical significance was set at the same threshold as in our primary analysis (i.e., two-sided *P*<0.05). We also evaluated which associations had effect estimates within the 95%CI of the venous thromboembolism-long COVID association.

### Gene-specific genetic instruments for venous thromboembolism

We constructed “gene-specific genetic instruments” by identifying variants associated with venous thromboembolism within or near different coagulation-related genes.

Coagulation-related genes were defined as those included in the “*Complement and coagulation cascades*” gene set from the KEGG database^72^ (**Supplementary Table 17**). Each gene-specific genetic instrument was constructed using genetic variants associated with venous thromboembolism within a *cis*-region 200 kilobases of the indicated coagulation-related gene in FinnGen^21^. Because a *cis*-region only comprises a small region within the genome, and because sensitivity analyses showed that genetic instruments for venous thromboembolism were still associated with long COVID when using more lenient *P*-value thresholds (i.e., *P*<5×10^-4^ or *P*<5×10^-6^) (**Supplementary Table 4**), we also included genetic variants associated with venous thromboembolism at a sub-genome-wide significance threshold to increase the number of genes with valid gene-specific genetic instruments. More specifically, if a gene’s *cis*-region included variants associated with venous thromboembolism at *P*<5×10^-8^, we used this threshold to construct the genetic instrument; if not, we used *P*<5×10^-6^ or *P*<5×10^-4^, depending on the presence of *cis*-variants surpassing those *P*-value thresholds. Independent genetic variants were selected using a linkage disequilibrium *R*^2^ threshold of 0.001.

MR analyses testing the effects of gene-specific genetic instruments with more than three variants used the inverse-variance weighted method with multiplicative random effects^12^; those instruments with two or three variants used the inverse-variance weighted method with fixed effects^12^; those testing the effects of a genetic instrument with only one variant used the Wald ratio estimator^65^. Considering that genetically predicted venous thromboembolism was strongly associated with long COVID and that this association was robust across multiple genetic instrument *P*-value thresholds, and that gene-specific genetic instruments inherently include fewer genetic variants than genetic instruments constructed using the entire genome, we did not formally correct these analyses for multiple comparisons. Instead, we prioritized associations based on effect size and used two-sided *P*<0.05 to indicate statistical significance.

### MR analyses using genetically predicted protein levels

Because *F2R*’s gene-specific genetic instrument had largest effect on long COVID across all gene-specific genetic instruments, downstream analyses focused on this gene and its gene products (i.e., *F2R* RNA and the PAR-1 protein). First, we tested the effects of genetically predicted circulating levels of PAR-1 on long COVID risk using a genetic instrument from the UK Biobank Pharma Proteomics Project (UKB-PPP)^37^.

The UKB-PPP is a collaboration between 13 biopharmaceutical companies that funded the measurement of 2,923 circulating proteins in participants from the UK Biobank, a population-based cohort of individuals aged 40–69 years recruited between 2006 and 2010 across the United Kingdom^37^. A total of 54,306 UK Biobank participants who donated blood samples at baseline or follow-up study visits were selected for proteomic profiling and were included in the UKB-PPP cohort. Blood samples were analyzed using proximity extension assay technology (Olink Explore 3072 platform [Olink Proteomics, Inc; Waltham, MA, USA]). Sun *et al.*^37^ performed a GWAS of circulating proteins measured using Olink technology in a discovery cohort of 35,571 UKB-PPP participants of European ancestry. Genotyping, imputation, and quality control of the UK Biobank samples were performed as described previously^37,73^. In addition to checking for sex mismatch, sex chromosome aneuploidy, and heterozygosity, imputed genetic variants were filtered for imputation INFO scoreD>0.7.

Individual protein levels were inverse-rank normalized prior to association analyses, which were adjusted for age, age^2^, sex, age × sex, age^2^ × sex, batch, assessment center, genetic array, time between blood sampling and measurement, and 20 principal components of genetic ancestry. The UK Biobank received approval from the North West Multi-center Research Ethics Committee, and all participants provided informed consent.

We constructed a genetic instrument for circulating PAR-1 levels using independent (linkage disequilibrium *R*^2^<0.001) genetic variants within 200 kilobases of *F2R* strongly associated with circulating PAR-1 levels (*P*<5×10^-8^). Such variants are also called *cis-*protein quantitative trait loci (*cis-*pQTLs) and have favorable effects on the assumptions of MR analyses^59–61^. The aforementioned thresholds resulted in only one variant (i.e., rs168753) to be included in the genetic instrument for circulating PAR-1; therefore, the association of this instrument with long COVID was tested using the Wald ratio estimator^65^. Sensitivity analyses used different *P*-value (*P*<5×10^-4^/*P*<5×10^-6^/*P*<5×10^-8^/*P*<5×10^-10^) and linkage disequilibrium *R*^2^ (*R*^2^<0.0001/*R*^2^<0.001/*R*^2^<0.01/*R*^2^<0.1) thresholds to construct genetic instruments. In addition, because *F2R*’s *cis*-region partially overlaps with those of neighboring genes (i.e., *F2RL1* and *F2RL2*)^42^, we also performed analyses with genetic instruments selectively including intragenic variants to limit pleiotropic effects of extragenic variants potentially influencing other genes’ expression patterns. Two-sided *P*<0.05 indicated statistical significance for this corroboratory analysis, and directional consistency was used as the criterium for robustness in sensitivity analyses.

### Colocalization analyses

To complement the MR, we performed colocalization analyses to test for shared causal variants between circulating PAR-1 levels and long COVID. Analyses were carried out with variants within 200 kilobases of *F2R*, using the *coloc.abf()* function from the R package *coloc* (version 5.2.2)^74^. The output of these analyses was expressed as test statistics that estimate the posterior probabilities of five hypotheses: (1) no causal variant for either trait; (2) a causal variant for the first but not the second trait; (3) a causal variant for the second but not the first trait; (4) distinct causal variants underlying both traits; and (5) one shared causal variant underlying both traits^38,39^. A high posterior probability for the fifth hypothesis (>80%) indicates strong evidence of colocalization, suggesting a high likelihood of a shared causal variant driving both traits. A high posterior probability for the fourth hypothesis (>80%) indicates strong evidence of distinct causal variants, suggesting a high likelihood of confounding by linkage disequilibrium (or horizontal pleiotropy) in the corresponding MR analysis. If there is no high probability for the fourth or fifth hypothesis (<80% for each) but the MR association is statistically significant and not a false positive finding, this suggests that the colocalization analysis is likely not adequately powered to distinguish between whether the MR association is due to a shared causal variant or a variant in linkage disequilibrium (i.e., horizontal pleiotropy)^38,39^. In the latter situation, we also estimated the probability for shared causal variants, conditional on there being at least one variant associated with each trait under study, calculated as: (PP*_H4_*) / (PP*_H3_* + PP*_H4_*)^38,39^, where PP*_H3_* indicates the posterior probability of the fourth hypothesis (i.e., distinct causal variants for both traits) and PP*_H4_* that of the fifth hypothesis (i.e., one shared causal variant for both traits).

### MR analyses using genetically predicted transcript levels

In addition to analyses focused on the PAR-1 protein, we also performed MR analyses testing the effects of genetically predicted *F2R* RNA levels on long COVID risk. To this end, we selected five tissues or cell types with (1) high *F2R* expression (queried using the Genome-Tissue Expression [GTEx] database^40^); (2) biological relevance to long COVID; and (3) at least one valid variant near *F2R* associated with *F2R* expression (i.e., *cis*- expression quantitative trait locus [*cis*-eQTL]). The highest median *F2R* RNA levels in GTEx were observed for the following tissues: aorta, coronary artery, tibial artery, skin (not sun-exposed), skin (sun-exposed), minor salivary gland, lung, and spleen^40^. Because we assumed that the skin (both sun-exposed and not sun-exposed) and minor salivary gland had less direct biological relevance to long COVID in comparison with the other tissues, we did not carry these tissues forward for further analyses. Given high *F2R* expression in platelets^34^—a cell type not included in the GTEx database^40^—we added platelets to our list of tissues and cell types of interest. Genetic instruments for platelets were obtained from the GeneSTAR Research Study^41^, while those for the other tissues (i.e., aorta, coronary artery, tibial artery, lung, and spleen) were queried in GTEx (v8)^40^.

We used the GTEx (v8) dataset^40^ to construct genetic instruments for *F2R* expression in the aforementioned tissues. In brief, the GTEx project has created a resource of gene expression levels from normal (i.e., non-diseased) tissues from deceased human donors.

GTEx included 838 donors with available RNA sequences and genotypes, collectively providing 17,382 samples from 52 tissues and two cell lines^75^. A total of 715 (85.3%) participants had European ancestry and were used to construct genetic instruments in the present analysis. RNA sequencing libraries were generated using the Illumina TruSeq protocol (Illumina Inc., San Diego, CA, USA); gene-level expression levels were quantified as reads per kilobase of transcript per million mapped reads^40^. Participants were genotyped using the Illumina Human Omni 2.5M and 5M Beadchips and imputed using the multi-ethnic reference panel from the 1000 Genomes Project (phase 3)^63^. Samples with fewer than 10 million mapped reads or outlier expression measurements were removed^40^. Variants with call rates <95%, minor allele frequencies <1%, Hardy-Weinberg equilibrium *P*<1×10^-6^, or imputation INFO score <0.4 were excluded. Genetic association analyses of transcript levels were adjusted for PEER factors, sex, genotyping platform, and three principal components of genetic ancestry. The protocol for the GTEx project was reviewed by Chesapeake Research Review Inc., Roswell Park Cancer Institute’s Office of Research Subject Protection, and the institutional review board of the University of Pennsylvania. For all donors, consent was obtained via next-of-kin consent^40^.

We used data from the GeneSTAR Research Study^41^ to construct genetic instruments for *F2R* expression in platelets. In brief, the GeneSTAR Research Study was designed to examine gene-environment determinants of platelet reactivity in response to low-dose aspirin therapy^76^. GeneSTAR included siblings identified from probands with coronary artery disease at age <60 years, as well as the spouses and adult offspring of the siblings. A total of 307 apparently healthy individuals were included for platelet eQTL analyses, of whom 180 (58.6%) had European ancestry and were used to construct genetic instruments in the present analysis^41^. Once platelets were isolated from whole blood samples, RNA sequencing libraries were generated using the Illumina TruSeq protocol (Illumina Inc., San Diego, CA, USA). Transcript abundances were quantified as fragments per kilobase of transcript per million sequenced reads. Participants underwent whole genome sequencing as part of the NHLBI’s Trans-Omics for Precision Medicine (TOPMed) program^77^. Low-expression outliers and samples with poor quality or unexpected familial relationships were excluded^41^. Variants were filtered for autosomal single-nucleotide polymorphisms with at least two samples per genotype and a call rate of >80%. Genetic association analyses of transcript levels were adjusted for sex, age, RNA sequencing batch, 15 principal components of the filtered and log-transformed gene expression matrix, and three principal components of genetic ancestry. The GeneSTAR Research Study was approved by the institutional review board of the Johns Hopkins Medical Institutions, and all participants provided informed consent^76^.

We constructed a genetic instrument for tissue-specific *F2R* expression using independent (linkage disequilibrium *R*^2^<0.001) genetic variants within 200 kilobases of *F2R* associated with *F2R* RNA levels from the GeneSTAR Research Study^41^ (for platelets) or GTEx project^40^ (for the other tissues). Similar to our analyses using gene-specific genetic instruments for venous thromboembolism (see **Gene-specific genetic instruments for venous thromboembolism**), each tissue-specific genetic instrument’s *P*-value threshold depended on the association strength of the different variants near *F2R*. If *F2R*’s *cis*-region included variants associated with tissue-specific *F2R* expression at *P*<5×10^-8^, we used this threshold to construct the genetic instrument; if not, we used *P*<5×10^-6^ or *P*<5×10^-4^, depending on the presence of *cis*-variants surpassing those *P*-value thresholds. This approach resulted in one genetic variant (i.e., the strongest *cis*-eQTL) for each tissue-specific genetic instrument; therefore, the association of these instruments with long COVID was tested using the Wald ratio estimator^65^. The only tissue without adequate *cis*-eQTLs for *F2R* was coronary arterial tissue, which was excluded from the analyses. Similar to the analyses using *cis-*pQTLs for PAR-1, we also performed analyses that only included intragenic variants. The genetic instruments for *F2R* expression in tibial arterial tissue included rs253072 (using variants within 200 kilobases) and rs250730 (using only intragenic variants), respectively; those for splenic tissue included rs253072 and rs250750; those for pulmonary tissue included rs2227750 and rs12735; and those for platelets included rs253072 and rs32934. We could only construct an adequate instrument for aortic tissue using variants within 200 kilobases (i.e., rs9293695) and not using intragenic variants alone. Two-sided *P*<0.05 indicated statistical significance for these confirmatory analyses.

### Data availability

This study used summary statistics from genome-wide association studies performed in FinnGen (release 10; available from https://r10.finngen.fi/), the Long COVID Host Genetics Initiative (available from https://my.locuszoom.org/gwas/793752/; https://doi.org/10.1101/2023.06.29.23292056), the Million Veteran Program (available from dbGAP, accession code no. phs001672.v2.p1), the PHOSP-COVID study (https://www.phosp.org/), the COVID-19 Host Genetics Initiative (release 7; available from https://www.covid19hg.org/results/r7/), the UK Biobank (available from http://ukb-ppp.gwas.eu/), the Genotype-Tissue Expression project (v10; available from https://gtexportal.org/home/downloads/adult-gtex/qtl), and the GeneSTAR Research Study (available from http://www.biostat.jhsph.edu/~kkammers/GeneSTAR/).

### Code availability

Code used for the main analyses of this study can be accessed at [link to be published upon publication].

## Data Availability

This study used summary statistics from genome-wide association studies performed in FinnGen (release 10; available from https://r10.finngen.fi/), the Long COVID Host Genetics Initiative (available from https://my.locuszoom.org/gwas/793752/; https://doi.org/10.1101/2023.06.29.23292056), the Million Veteran Program (available from dbGAP, accession code no. phs001672.v2.p1), the PHOSP-COVID study (https://www.phosp.org/), the COVID-19 Host Genetics Initiative (release 7; available from https://www.covid19hg.org/results/r7/), the UK Biobank (available from http://ukb-ppp.gwas.eu/), the Genotype-Tissue Expression project (v10; available from https://gtexportal.org/home/downloads/adult-gtex/qtl), and the GeneSTAR Research Study (available from http://www.biostat.jhsph.edu/∼kkammers/GeneSTAR/).

https://my.locuszoom.org/gwas/793752/

https://doi.org/10.1101/2023.06.29.23292056

https://www.phosp.org/

https://www.covid19hg.org/results/r7/

http://ukb-ppp.gwas.eu/

https://gtexportal.org/home/downloads/adult-gtex/qtl

http://www.biostat.jhsph.edu/~kkammers/GeneSTAR/

## Acknowledgements

The authors acknowledge the participants and investigators of the FinnGen study, the Long COVID Host Genetics Initiative, the Million Veteran Program, the Post-Hospitalization COVID-19 study, the UK Biobank, the Genotype-Tissue Expression project, and the GeneSTAR Research Study. A.S. is supported by Life Sciences Research Partners (LSRP).

T.N. is supported by a research fellowship of the Japan Society for the Promotion of Science for Young Scientists (22J30004) and by a Grant-in-Aid for Scientific Research (B) (23H02917). M.K.G. is funded by the German Research Foundation in the form of an Emmy Noether grant (GZ: GE 3461/2-1, ID 512461526). B.G.G. is supported by the Wellcome Trust (221680/Z/20/Z). L.V.W. holds a GlaxoSmithKline Asthma + Lung UK Chair in Respiratory Research (C17-1). B.R. is supported by a Wellcome Career Development Award fellowship (302210/Z/23/Z). H.Z. is supported by the Swedish Research Council (2021-03050), the Knut and Alice Wallenberg Foundation (2023.0141), Groschinskys Minnesfond, the Max Planck Society as well as Cornell’s, Philip-Sörensen’s, and Hedlund’s foundations. H.M.O. is supported by the Academy of Finland (#1350181) and the NIH (R01AI170850). K.F. is supported by the National Institute of Health (R01Hl161365), KU Leuven (C14/23/121), research foundation Flanders (FWO; G072921N). T.V. is supported by a grant from FWO (1843423N). PHOSP-COVID is jointly funded by a grant from the MRC-UK Research and Innovation and the Department of Health and Social Care through the National Institute for Health Research (NIHR) rapid response panel to tackle COVID-19 (grant references: MR/V027859/1 and COV0319). The views expressed in the publication are those of the author(s) and not necessarily those of the National Health Service (NHS), the NIHR or the Department of Health and Social Care. This study would not be possible without all the participants who have given their time and support. We thank all the participants and their families. We thank the many research administrators, health-care and social-care professionals who contributed to setting up and delivering the study at all of the 65 NHS trusts/Health boards and 25 research institutions across the UK, as well as all the supporting staff at the NIHR Clinical Research Network, Health Research Authority, Research Ethics Committee, Department of Health and Social Care, Public Health Scotland, and Public Health England, and support from the ISARIC Coronavirus Clinical Characterisation Consortium. We thank Kate Holmes at the NIHR Office for Clinical Research Infrastructure (NOCRI) for her support in coordinating the charities group. The PHOSP-COVID industry framework was formed to provide advice and support in commercial discussions, and we thank the Association of the British Pharmaceutical Industry as well NOCRI for coordinating this. We are very grateful to all the charities that have provided insight to the study: Action Pulmonary Fibrosis, Alzheimer’s Research UK, Asthma + Lung UK, British Heart Foundation, Diabetes UK, Cystic Fibrosis Trust, Kidney Research UK, MQ Mental Health, Muscular Dystrophy UK, Stroke Association Blood Cancer UK, McPin Foundations, and Versus Arthritis. We thank the NIHR Leicester Biomedical Research Centre patient and public involvement group and Long Covid Support.

## Inclusion and ethics

Inclusion and ethics standards have been reviewed where applicable.

## Corresponding authors

Correspondence and requests for materials should be addressed to A.S.

## References

1. Vos T., Hanson SW., Abbafati C., Aerts JG., Al-Aly Z., Ashbaugh C., et al. Estimated Global Proportions of Individuals With Persistent Fatigue, Cognitive, and Respiratory Symptom Clusters Following Symptomatic COVID-19 in 2020 and 2021. JAMA 2022;328:1604–15. 10.1001/JAMA.2022.18931.

2. Soriano JB., Murthy S., Marshall JC., Relan P., Diaz J V. A clinical case definition of post-COVID-19 condition by a Delphi consensus. Lancet Infect Dis 2022;22:e102–7. 10.1016/S1473-3099(21)00703-9.

3. Davis HE., McCorkell L., Vogel JM., Topol EJ. Long COVID: major findings, mechanisms and recommendations. Nat Rev Microbiol 2023;21:133–46. 10.1038/s41579-022-00846-2.

4. Seeble J., Waterboer T., Hippchen T., Simon J., Kirchner M., Lim A., et al. Persistent Symptoms in Adult Patients 1 Year After Coronavirus Disease 2019 (COVID-19): A Prospective Cohort Study. Clin Infect Dis 2022;74:1191–8. 10.1093/CID/CIAB611.

5. Tran VT., Porcher R., Pane I., Ravaud P. Course of post COVID-19 disease symptoms over time in the ComPaRe long COVID prospective e-cohort. Nat Commun 2022;13:1–6. 10.1038/s41467-022-29513-z.

6. Taquet M., Skorniewska Z., Hampshire A., Chalmers JD., Ho LP., Horsley A., et al. Acute blood biomarker profiles predict cognitive deficits 6 and 12 months after COVID-19 hospitalization. Nat Med 2023;29:2498–508. 10.1038/s41591-023-02525-y.

7. Cervia-Hasler C., Brüningk SC., Hoch T., Fan B., Muzio G., Thompson RC., et al. Persistent complement dysregulation with signs of thromboinflammation in active Long Covid. Science (1979) 2024;383:1–18. 10.1126/SCIENCE.ADG7942.

8. Davies M. Long covid patients travel abroad for expensive and experimental “blood washing.” BMJ 2022;378. 10.1136/BMJ.O1671.

9. Uffelmann E., Huang QQ., Munung NS., de Vries J., Okada Y., Martin AR., et al. Genome-wide association studies. Nat Rev Methods Primers 2021;1:1–21. 10.1038/s43586-021-00056-9.

10. Davies NM., Holmes M V., Davey Smith G. Reading Mendelian randomisation studies: a guide, glossary, and checklist for clinicians. BMJ 2018;362:601. 10.1136/BMJ.K601.

11. Sanderson E., Glymour MM., Holmes M V., Kang H., Morrison J., Munafò MR., et al. Mendelian randomization. Nat Rev Methods Primers 2022;2:1–21. 10.1038/s43586-021-00092-5.

12. Burgess S., Butterworth A., Thompson SG. Mendelian Randomization Analysis With Multiple Genetic Variants Using Summarized Data. Genet Epidemiol 2013;37:658–65. 10.1002/GEPI.21758.

13. Ardissino M., Morley AP., Slob EAW., Schuermans A., Rayes B., Raisi-Estabragh Z., et al. Birth weight influences cardiac structure, function, and disease risk: evidence of a causal association. Eur Heart J 2023. 10.1093/EURHEARTJ/EHAD631.

14. Gaziano L., Giambartolomei C., Pereira AC., Gaulton A., Posner DC., Swanson SA., et al. Actionable druggable genome-wide Mendelian randomization identifies repurposing opportunities for COVID-19. Nat Med 2021;27:668–76. 10.1038/s41591-021-01310-z.

15. Bovijn J., Lindgren CM., Holmes M V. Genetic variants mimicking therapeutic inhibition of IL-6 receptor signaling and risk of COVID-19. Lancet Rheumatol 2020;2:e658–9. 10.1016/S2665-9913(20)30345-3.

16. Niemi MEK., Karjalainen J., Liao RG., Neale BM., Daly M., Ganna A., et al. Mapping the human genetic architecture of COVID-19. Nature 2021;600:472–7. 10.1038/s41586-021-03767-x.

17. 17. Interleukin-6 Receptor Antagonists in Critically Ill Patients with Covid-19. N Engl J Med 2021;384:1491–502. 10.1056/NEJMOA2100433.

18. 18. Group TWREA for C-19 T (REACT) W., Domingo P., Mur I., Mateo GM., Gutierrez M del M., Pomar V., et al. Association Between Administration of IL-6 Antagonists and Mortality Among Patients Hospitalized for COVID-19: A Meta-analysis. JAMA 2021;326:499–518. 10.1001/JAMA.2021.11330.

19. Trajanoska K., Bhérer C., Taliun D., Zhou S., Richards JB., Mooser V. From target discovery to clinical drug development with human genetics. Nature 2023;620:737–45. 10.1038/s41586-023-06388-8.

20. Pierce BL., Burgess S. Efficient Design for Mendelian Randomization Studies: Subsample and 2-Sample Instrumental Variable Estimators. Am J Epidemiol 2013;178:1177–84. 10.1093/AJE/KWT084.

21. Kurki MI., Karjalainen J., Palta P., Sipilä TP., Kristiansson K., Donner KM., et al. FinnGen provides genetic insights from a well-phenotyped isolated population. Nature 2023;613:508–18. 10.1038/s41586-022-05473-8.

22. Leinonen JT., Pirinen M., Tukiainen T. Disentangling the link between maternal influences on birth weight and disease risk in 36,211 genotyped mother–child pairs. Commun Biol 2024;7:1–9. 10.1038/s42003-024-05872-9.

23. Access results | FinnGen. Available at: https://www.finngen.fi/en/access_results. Accessed October 24, 2024.

24. Lammi V., Nakanishi T., Jones SE., Andrews SJ., Karjalainen J., Cortés B., et al. Genome-wide Association Study of Long COVID. MedRxiv. 10.1101/2023.06.29.23292056.

25. Burgess S., Thompson SG. Interpreting findings from Mendelian randomization using the MR-Egger method. Eur J Epidemiol 2017;32:377. 10.1007/S10654-017-0255-X.

26. Hemani G., Tilling K., Davey Smith G. Orienting the causal relationship between imprecisely measured traits using GWAS summary data. PLoS Genet 2017;13:e1007081. 10.1371/JOURNAL.PGEN.1007081.

27. Klarin D., Busenkell E., Judy R., Lynch J., Levin M., Haessler J., et al. Genome-wide association analysis of venous thromboembolism identifies new risk loci and genetic overlap with arterial vascular disease. Nat Genet 2019;51:1574–9. 10.1038/s41588-019-0519-3.

28. Evans RA., McAuley H., Harrison EM., Shikotra A., Singapuri A., Sereno M., et al. Physical, cognitive, and mental health impacts of COVID-19 after hospitalisation (PHOSP-COVID): a UK multicentre, prospective cohort study. Lancet Respir Med 2021;9:1275–87. 10.1016/S2213-2600(21)00383-0.

29. Elneima O., McAuley HJC., Leavy OC., Chalmers JD., Horsley A., Ho L-P., et al. Cohort Profile: Post-Hospitalisation COVID-19 (PHOSP-COVID) study. Int J Epidemiol 2024;53:165. 10.1093/IJE/DYAD165.

30. Bilaloglu S., Aphinyanaphongs Y., Jones S., Iturrate E., Hochman J., Berger JS. Thrombosis in Hospitalized Patients With COVID-19 in a New York City Health System. JAMA 2020;324:799–801. 10.1001/JAMA.2020.13372.

31. Katsoularis I., Fonseca-Rodríguez O., Farrington P., Jerndal H., Lundevaller EH., Sund M., et al. Risks of deep vein thrombosis, pulmonary embolism, and bleeding after covid-19: nationwide self-controlled cases series and matched cohort study. BMJ 2022;377:e069590. 10.1136/BMJ-2021-069590.

32. Kanai M., Andrews SJ., Cordioli M., Stevens C., Neale BM., Daly M., et al. A second update on mapping the human genetic architecture of COVID-19. Nature 2023;621:E7–26. 10.1038/s41586-023-06355-3.

33. Burgess S., Thompson SG. Multivariable Mendelian Randomization: The Use of Pleiotropic Genetic Variants to Estimate Causal Effects. Am J Epidemiol 2015;181:251–60. 10.1093/AJE/KWU283.

34. Angiolillo DJ., Capodanno D., Goto S. Platelet thrombin receptor antagonism and atherothrombosis. Eur Heart J 2010;31:17–28. 10.1093/EURHEARTJ/EHP504.

35. Smith SMG., Judge HM., Peters G., Armstrong M., Dupont A., Gaussem P., et al. PAR-1 genotype influences platelet aggregation and procoagulant responses in patients with coronary artery disease prior to and during clopidogrel therapy. Platelets 2005;16:340–5. 10.1080/00207230500120294.

36. Dupont A., Fontana P., Bachelot-Loza C., Reny JL., Bièche I., Desvard F., et al. An intronic polymorphism in the PAR-1 gene is associated with platelet receptor density and the response to SFLLRN. Blood 2003;101:1833–40. 10.1182/BLOOD-2002-07-2149.

37. Sun BB., Chiou J., Traylor M., Benner C., Hsu YH., Richardson TG., et al. Plasma proteomic associations with genetics and health in the UK Biobank. Nature 2023;622:329–38. 10.1038/s41586-023-06592-6.

38. Woolf B., Rajasundaram S., Cronjé HT., Yarmolinsky J., Burgess S., Gill D. A drug target for erectile dysfunction to help improve fertility, sexual activity, and wellbeing: mendelian randomisation study. BMJ 2023;383. 10.1136/BMJ-2023-076197.

39. Zuber V., Grinberg NF., Gill D., Manipur I., Slob EAW., Patel A., et al. Combining evidence from Mendelian randomization and colocalization: Review and comparison of approaches. Am J Hum Genet 2022;109:767–82. 10.1016/J.AJHG.2022.04.001/ATTACHMENT/FC339381-C46B-413F-A624-9A1B10820BED/MMC1.PDF.

40. Aguet F., Brown AA., Castel SE., Davis JR., He Y., Jo B., et al. Genetic effects on gene expression across human tissues. Nature 2017;550:204–13. 10.1038/nature24277.

41. Kammers K., Taub MA., Rodriguez B., Yanek LR., Ruczinski I., Martin J., et al. Transcriptional profile of platelets and iPSC-derived megakaryocytes from whole-genome and RNA sequencing. Blood 2021;137:959–68. 10.1182/BLOOD.2020006115.

42. Kahn ML., Hammes SR., Botka C., Coughlin SR. Gene and Locus Structure and Chromosomal Localization of the Protease-activated Receptor Gene Family. J Biol Chem 1998;273:23290–6. 10.1074/JBC.273.36.23290.

43. Pretorius E., Vlok M., Venter C., Bezuidenhout JA., Laubscher GJ., Steenkamp J., et al. Persistent clotting protein pathology in Long COVID/Post-Acute Sequelae of COVID-19 (PASC) is accompanied by increased levels of antiplasmin. Cardiovasc Diabetol 2021;20:1–18. 10.1186/S12933-021-01359-7/FIGURES/7.

44. Klok FA., Kruip MJHA., van der Meer NJM., Arbous MS., Gommers DAMPJ., Kant KM., et al. Incidence of thrombotic complications in critically ill ICU patients with COVID-19. Thromb Res 2020;191:145–7. 10.1016/J.THROMRES.2020.04.013.

45. Conway EM., Mackman N., Warren RQ., Wolberg AS., Mosnier LO., Campbell RA., et al. Understanding COVID-19-associated coagulopathy. Nat Rev Immunol 2022;22:639–49. 10.1038/s41577-022-00762-9.

46. Kanth Manne B., Denorme F., Middleton EA., Portier I., Rowley JW., Stubben C., et al. Platelet gene expression and function in patients with COVID-19. Blood 2020;136:1317–29. 10.1182/BLOOD.2020007214.

47. Bryce C., Grimes Z., Pujadas E., Ahuja S., Beasley MB., Albrecht R., et al. Pathophysiology of SARS-CoV-2: the Mount Sinai COVID-19 autopsy experience. Mod Pathol 2021;34:1456–67. 10.1038/s41379-021-00793-y.

48. Ackermann M., Verleden SE., Kuehnel M., Haverich A., Welte T., Laenger F., et al. Pulmonary Vascular Endothelialitis, Thrombosis, and Angiogenesis in Covid-19. N Engl J Med 2020;383:120–8. 10.1056/NEJMOA2015432.

49. Oxley TJ., Mocco J., Majidi S., Kellner CP., Shoirah H., Singh IP., et al. Large-Vessel Stroke as a Presenting Feature of Covid-19 in the Young. N Engl J Med 2020;382. 10.1056/NEJMC2009787.

50. Stevens H., Canovas R., Tran H., Peter K., McFadyen JD. Inherited Thrombophilias Are Associated With a Higher Risk of COVID-19–Associated Venous Thromboembolism: A Prospective Population-Based Cohort Study. Circulation 2022;145:940–2. 10.1161/CIRCULATIONAHA.121.057394.

51. Ryu JK., Yan Z., Montano M., Sozmen EG., Dixit K., Suryawanshi RK., et al. Fibrin drives thromboinflammation and neuropathology in COVID-19. Nature 2024;633:905–13. 10.1038/s41586-024-07873-4.

52. Swank Z., Senussi Y., Manickas-Hill Z., Yu XG., Li JZ., Alter G., et al. Persistent Circulating Severe Acute Respiratory Syndrome Coronavirus 2 Spike Is Associated With Post-acute Coronavirus Disease 2019 Sequelae. Clinical Infectious Diseases 2023;76:e487–90. 10.1093/CID/CIAC722.

53. Coughlin SR. Thrombin signalling and protease-activated receptors. Nature 2000;407:258–64. 10.1038/35025229.

54. Morrow DA., Braunwald E., Bonaca MP., Ameriso SF., Dalby AJ., Fish MP., et al. Vorapaxar in the Secondary Prevention of Atherothrombotic Events. N Engl J Med 2012;366:1404–13. 10.1056/NEJMOA1200933.

55. Tricoci P., Huang Z., Held C., Moliterno DJ., Armstrong PW., Van de Werf F., et al. Thrombin-Receptor Antagonist Vorapaxar in Acute Coronary Syndromes. N Engl J Med 2012;366:20–33. 10.1056/NEJMOA1109719/SUPPL_FILE/NEJMOA1109719_DISCLOS URES.PDF.

56. Niessen F., Schaffner F., Furlan-Freguia C., Pawlinski R., Bhattacharjee G., Chun J., et al. Dendritic cell PAR1–S1P3 signalling couples coagulation and inflammation. Nature 2008;452:654–8. 10.1038/nature06663.

57. Subramaniam S., Ogoti Y., Hernandez I., Zogg M., Botros F., Burns R., et al. A thrombin-PAR1/2 feedback loop amplifies thromboinflammatory endothelial responses to the viral RNA analogue poly(I:C). Blood Adv 2021;5:2760–74. 10.1182/BLOODADVANCES.2021004360.

58. Burgess S., Davies NM., Thompson SG. Bias due to participant overlap in two-sample Mendelian randomization. Genet Epidemiol 2016;40:597–608. 10.1002/GEPI.21998.

59. Schuermans A., Truong B., Ardissino M., Bhukar R., Slob EAW., Nakao T., et al. Genetic Associations of Circulating Cardiovascular Proteins With Gestational Hypertension and Preeclampsia. JAMA Cardiol 2024. 10.1001/JAMACARDIO.2023.4994.

60. Schuermans A., Pournamdari AB., Lee J., Bhukar R., Ganesh S., Darosa N., et al. Integrative proteomic analyses across common cardiac diseases yield mechanistic insights and enhanced prediction. Nature Cardiovascular Research 2024:1–15. 10.1038/s44161-024-00567-0.

61. Swerdlow DI., Kuchenbaecker KB., Shah S., Sofat R., Holmes M V., White J., et al. Selecting instruments for Mendelian randomization in the wake of genome-wide association studies. Int J Epidemiol 2016;45:1600–16. 10.1093/IJE/DYW088.

62. FinnGen Public Documentation. Available at: https://finngen.gitbook.io/documentation/r10. Accessed November 5, 2024.

63. The 1000 Genomes Project Consortium. A global reference for human genetic variation. Nature 2015;526:68–74. 10.1038/nature15393.

64. Purcell S., Neale B., Todd-Brown K., Thomas L., Ferreira MAR., Bender D., et al. PLINK: A tool set for whole-genome association and population-based linkage analyses. Am J Hum Genet 2007;81:559–75. 10.1086/519795.

65. Wald A. The Fitting of Straight Lines if Both Variables are Subject to Error. Ann Math Stat 1940;11:284–300. 10.1214/AOMS/1177731868.

66. Hemani G., Zheng J., Elsworth B., Wade KH., Haberland V., Baird D., et al. The MR-base platform supports systematic causal inference across the human phenome. Elife 2018;7. 10.7554/ELIFE.34408.

67. Yavorska OO., Burgess S. MendelianRandomization: an R package for performing Mendelian randomization analyses using summarized data. Int J Epidemiol 2017;46:1734–9. 10.1093/IJE/DYX034.

68. Bowden J., Davey Smith G., Haycock PC., Burgess S. Consistent Estimation in Mendelian Randomization with Some Invalid Instruments Using a Weighted Median Estimator. Genet Epidemiol 2016;40:304–14. 10.1002/GEPI.21965.

69. Hartwig FP., Smith GD., Bowden J. Robust inference in summary data Mendelian randomization via the zero modal pleiotropy assumption. Int J Epidemiol 2017;46:1985–98. 10.1093/IJE/DYX102.

70. Henry A., Gordillo-Marañón M., Finan C., Schmidt AF., Ferreira JP., Karra R., et al. Therapeutic Targets for Heart Failure Identified Using Proteomics and Mendelian Randomization. Circulation 2022;145:1205–17. 10.1161/CIRCULATIONAHA.121.056663.

71. Gaziano JM., Concato J., Brophy M., Fiore L., Pyarajan S., Breeling J., et al. Million Veteran Program: A mega-biobank to study genetic influences on health and disease. J Clin Epidemiol 2016;70:214–23. 10.1016/J.JCLINEPI.2015.09.016.

72. Kanehisa M., Goto S. KEGG: Kyoto Encyclopedia of Genes and Genomes. Nucleic Acids Res 2000;28:27–30. 10.1093/NAR/28.1.27.

73. Bycroft C., Freeman C., Petkova D., Band G., Elliott LT., Sharp K., et al. The UK Biobank resource with deep phenotyping and genomic data. Nature 2018;562:203–9. 10.1038/s41586-018-0579-z.

74. Giambartolomei C., Vukcevic D., Schadt EE., Franke L., Hingorani AD., Wallace C., et al. Bayesian Test for Colocalisation between Pairs of Genetic Association Studies Using Summary Statistics. PLoS Genet 2014;10:e1004383. 10.1371/JOURNAL.PGEN.1004383.

75. Aguet F., Barbeira AN., Bonazzola R., Brown A., Castel SE., Jo B., et al. The GTEx Consortium atlas of genetic regulatory effects across human tissues. Science (1979) 2020;369:1318–30. 10.1126/SCIENCE.AAZ1776/SUPPL_FILE/AAZ1776_TABLESS10-S16.XLSX.

76. Becker DM., Segal J., Vaidya D., Yanek LR., Herrera-Galeano JE., Bray PF., et al. Sex Differences in Platelet Reactivity and Response to Low-Dose Aspirin Therapy. JAMA 2006;295:1420–7. 10.1001/JAMA.295.12.1420.

77. Taliun D., Harris DN., Kessler MD., Carlson J., Szpiech ZA., Torres R., et al. Sequencing of 53,831 diverse genomes from the NHLBI TOPMed Program. Nature 2021;590:290–9. 10.1038/s41586-021-03205-y.

